# Disagreement between AI and Radiologists: Quantifying Direction and Fragility

**DOI:** 10.64898/2026.10.19.26364791

**Authors:** Bilal El Kaddouri, Pierre Alain Gevenois, Alexander A. Bankier

## Abstract

**Background:** Conventional performance metrics do not indicate whether artificial intelligence (AI)-platforms and radiologists are right or wrong on the same examinations, nor whether their non-agreement is more often beneficial or harmful.

**Objective:** To develop, illustrate, and apply directional metrics quantifying non-agreement between radiologists and AI-platforms against a common reference standard, and to adapt a fragility-based framework for quantifying the statistical robustness of these metrics.

**Methods:** We defined Positive Non-Agreement (PNA, AI-platform correct, radiologist incorrect), Negative Non-Agreement (NNA, radiologist correct, AI-platform incorrect), and Net Non-Agreement (NENA, their difference). Metrics were illustrated in three hypothetical scenarios of 1,000 examinations each and applied to three previously published AI-radiologist studies on pulmonary embolism and mammography. Bootstrap confidence intervals were calculated, and a reclassification fragility index (FI) quantified the minimum case reclassification needed to remove statistical support for NENA’s direction.

**Results:** In illustration, NENA was positive in two scenarios and negative in one. In the three published studies, NENA ranged from −4.18% to +10.74% and decreased consistently with increasing radiologist experience (junior residents, +6.27%; consultants, +1.45%), a gradient driven primarily by declining PNA with modest increase in NNA. The reclassification FI was defined for all three illustrative scenarios (range, 0.10%-1.70%) and for six of the seven comparisons from these studies (range, 0.34%-4.31%), but not for the weakest comparison, whose confidence interval for NENA included zero.

**Conclusions:** PNA, NNA, and NENA provide a directional, case-level complement to conventional performance metrics for AI-radiologist non-agreement. The accompanying reclassification FI offers a statistically grounded, generalizable method for quantifying their robustness.

**Summary Sentence:** Positive, Negative, and Net Non-Agreement, directional metrics developed and applied to published datasets comparing artificial intelligence with radiologists, show that the net benefit of artificial intelligence assistance depends on reader experience and clinical setting, and that the statistical robustness of these directional conclusions varies by comparison and sample size, which is information that conventional performance metrics alone do not provide.

**Key Points:**

- Positive Non-Agreement (PNA), Negative Non-Agreement (NNA), and Net Non-Agreement (NENA) are directional metrics that identify whether AI-radiologist non-agreement is more likely to be beneficial or harmful, complementing conventional performance metrics to inform AI validation and monitoring decisions.
- The reclassification fragility index (FI), anchored to statistical significance rather than to the point estimate, quantifies how many case reclassifications would remove support for the direction of NENA.
- Applied to three previously published AI-radiologist datasets NENA ranged from −4.18% to +10.74% and fell from +6.27% to +1.45% with increasing radiologist experience, a gradient driven primarily by declining PNA with modest increase in NNA.

## Introduction

Conventional performance metrics, such as sensitivity, specificity, accuracy, and predictive values, quantify how diagnostic tests perform overall, as compared to an independent reference standard (1, 2). Comparisons between radiologists and artificial intelligence (AI)-platforms often report aggregate accuracy for entire cohorts (3–7). However, little is known about comparisons on the same individual examinations, and no method currently exists to characterize potential non-agreement between radiologists and AI-platforms in this setting. This gap has consequences for how an AI-platform is deployed. Two clinical settings can report an identical improvement in AI-platform accuracy, while differing in what improvement consists of. In one, nearly all of it may come from the AI-platform correcting radiologist errors. In another, a comparable share may come from the AI-platform introducing errors a radiologist would otherwise not have made. Aggregate accuracy cannot distinguish between these situations, yet the distinction determines whether an AI-platform can reasonably operate with reduced oversight. When radiologists and AI-platforms disagree on the same examination compared with a common reference standard, either the radiologist or the AI-platform will be “right” and the other “wrong”: if the AI-platform is right, it corrects an erroneous decision and is beneficial; if it is wrong, it introduces an error and is harmful. No existing method separates these two types of non-agreement. This information, however, is important for local validation, quality and safety surveillance, and evaluation of automation bias (8–11). It could also help explain why reported AI-platform contributions vary with radiologist experience and institutional framework (3, 12). Because any interpretation of such an approach will depend on both the reference standard’s robustness and the statistical uncertainty of a finite sample, a complete framework should also indicate how much perturbation, from either source, would overturn it. Therefore, the aim of our study was to develop, illustrate, and test the applicability of a set of metrics to quantify non-agreement between radiologists and AI-platforms on the same individual examinations, using a common reference standard.

## Materials and Methods

### Development

In most studies, individual imaging examinations are interpreted independently by radiologists and AI-platforms, and each interpretation is compared to a common reference standard. Four mutually exclusive outcomes are possible: 1) both radiologist and AI-platform are right; 2) both are wrong; 3) the AI-platform is right and the radiologist is wrong; or 4) the radiologist is right and the AI-platform is wrong. The last two outcomes represent two distinct non-agreement patterns: AI-platform right and radiologist wrong (Positive Non-Agreement, PNA); and radiologist right and AI-platform wrong (Negative Non-Agreement, NNA). Here, “positive” and “negative” refer to the direction of non-agreement relative to the reference standard; they do not denote disease status. Net Non-Agreement (NENA) is the difference between PNA and NNA (Table 1). This paired-difference structure is conceptually related to classic approaches for comparing proportions, such as the McNemar test, although PNA and NNA additionally decompose the direction of paired discordance (13). Both PNA and NNA range from 0 to 1 (0% to 100%): a PNA value of 0 indicates that there is no examination in which the AI-platform is correct and the radiologist is wrong, and a PNA value of 1 indicates that this is the case for all examinations; the same logic applies to NNA, with the roles of the AI-platform and the radiologist reversed. NENA ranges from −1 to +1 (−100% to +100%), being positive when PNA occurs more often than NNA and negative when the opposite occurs. A NENA value of 0 indicates that PNA and NNA are equal, which does not necessarily represent perfect agreement, as it can reflect either the complete absence of non-agreement or equal and canceling numbers of PNA and NNA events.

**Table 1.** Directional paired-reader framework for calculation of Positive Non-Agreement (PNA), Negative Non-Agreement (NNA), and Net Non-Agreement (NENA). Each examination is classified according to whether the AI model and the radiologist are either concordant or discordant with the reference standard. Examinations for which the AI model is reference-standard concordant, and the radiologist is reference-standard discordant are classified as positive non-agreement (PNA) events. Cases in which the radiologist is reference-standard concordant, and the AI model is reference-standard discordant are classified as negative non-agreement (NNA) events. PNA and NNA quantify the frequency of these two directional non-agreement patterns, and net non-agreement (NENA) reflects the difference between PNA and NNA.

|  | *Reader correct* | *Reader incorrect* |
| --- | --- | --- |
| *AI correct* | AI and reader agree | AI is correct, reader is incorrect<br>→ *positive non-agreement event (PNA)* |
| *AI incorrect* | Reader is correct, AI is incorrect<br>→ *negative non-agreement (NNA)* | AI and reader agree |
| ***PNA-NNA = net non-agreement (NENA)*** |  |  |

### Illustration

The purpose of this illustration was to evaluate the feasibility of PNA, NNA, and NENA computations, and to compare their informative values with those of conventional performance metrics. We assumed an illustrative cohort of 1,000 cases, with 400 defined as disease-positive and 600 as disease-negative, and designed three scenarios: scenario 1, with positive NENA and low NNA; scenario 2, with positive NENA and high NNA; and scenario 3, with negative NENA and high NNA. The choice of numbers underlying PNAs and NNAs was arbitrary and the number of scenarios was limited to three for practical reasons. For each scenario, we also calculated the 95% bootstrap confidence interval for NENA and the reclassification fragility index (FI), which quantifies how many case reclassifications would remove statistical support for the direction of NENA (see Statistics). PNA, NNA, NENA, and conventional performance metrics are proportions that do not depend on cohort size, whereas their confidence intervals and the reclassification FI do; a cohort size of 1,000 was chosen so that the statistical robustness of each scenario could also be illustrated (Appendix A5).

### Applicability

To determine the relevance of the metric-triplet in a clinical context, we selected three studies from the literature to which these metrics could be applied. Here, applicability means that the metric-triplet yields plausible and clinically interpretable results when applied to real paired AI-radiologist data; it does not imply generalizability to other cohorts. As the selection of these three studies was not based on a systematic review, other eligible studies may exist. Studies were required to report, in the same individual examinations, radiologist-alone and AI-platform-alone assessments compared to a common reference standard, with sufficient data to reconstruct the counts defined below. We selected three studies that met these data requirements (5–7). We extracted the total number of examinations (Q), the number of PNA events (P; radiologist wrong, AI-platform right), and the number of NNA events (N; radiologist right, AI-platform wrong), which were used to calculate the metric-triplet as follows: PNA=P/Q; NNA=N/Q; and NENA=PNA-NNA (or (P-N)/Q). In addition, we computed sensitivity, specificity, PPV, NPV, and accuracy ourselves when not reported. As for the illustrative scenarios, we calculated the 95% bootstrap confidence interval for NENA and the reclassification FI for each comparison.

### Statistics

Confidence intervals for PNA, NNA, and NENA were determined using multinomial bootstrap resampling based on the observed counts of PNA, NNA, and concordant assessments (14–16). The null hypothesis of “no net directional effect” was evaluated by testing whether this interval included zero.

Detailed notations and calculation codes are given in Appendices A1 and A7.

To ensure the reclassification FI conveys information beyond NENA, we anchored it to statistical significance rather than the point estimate, since a point-estimate-based count is a fixed rescaling of NENA and adds no independent information (Appendix A1). We defined the reclassification FI, by analogy with the Fragility Index used in randomized trials (17), as the minimum number of case reclassifications required for the 95% bootstrap CI for NENA to include zero. In brief, it answers the question “how many examinations would need to be reclassified before we could no longer be confident an AI-platform’s net effect is beneficial or harmful, rather than indistinguishable from chance?”. If the baseline 95% confidence interval for NENA already included zero, the reclassification FI was not applicable, mirroring the convention that a trial-level Fragility Index is undefined for a non-significant trial. This procedure was applied to both the illustrative scenarios and the applicability studies. For datasets with multiple reader assessments of the same examinations, the aggregate published data did not permit adjustment for within-examination clustering. The full procedure and a worked example are detailed in Appendix A1. All statistical analyses were performed using R (version 4.6.1, R Foundation for Statistical Computing, Vienna, Austria).

## Results

### Development and Illustration

The application of our approach to three clinical scenarios is summarized in Table 2. NENA was positive in scenarios 1 and 2, and negative in scenario 3, despite moderate sensitivity and high specificity. AI-platform sensitivity was fixed at 80.0% across all three scenarios, whereas radiologist sensitivity ranged from 65.0% to 72.5%. Despite this constant AI-platform sensitivity, NNA varied more than ten-fold, from 1% in scenario 1 to 15% in scenario 2, and specificity showed no consistent relationship to the direction or magnitude of NNA or NENA across scenarios. Confidence intervals of the metrics in Table 2 are reported in Appendix A5.

**Table 2.**
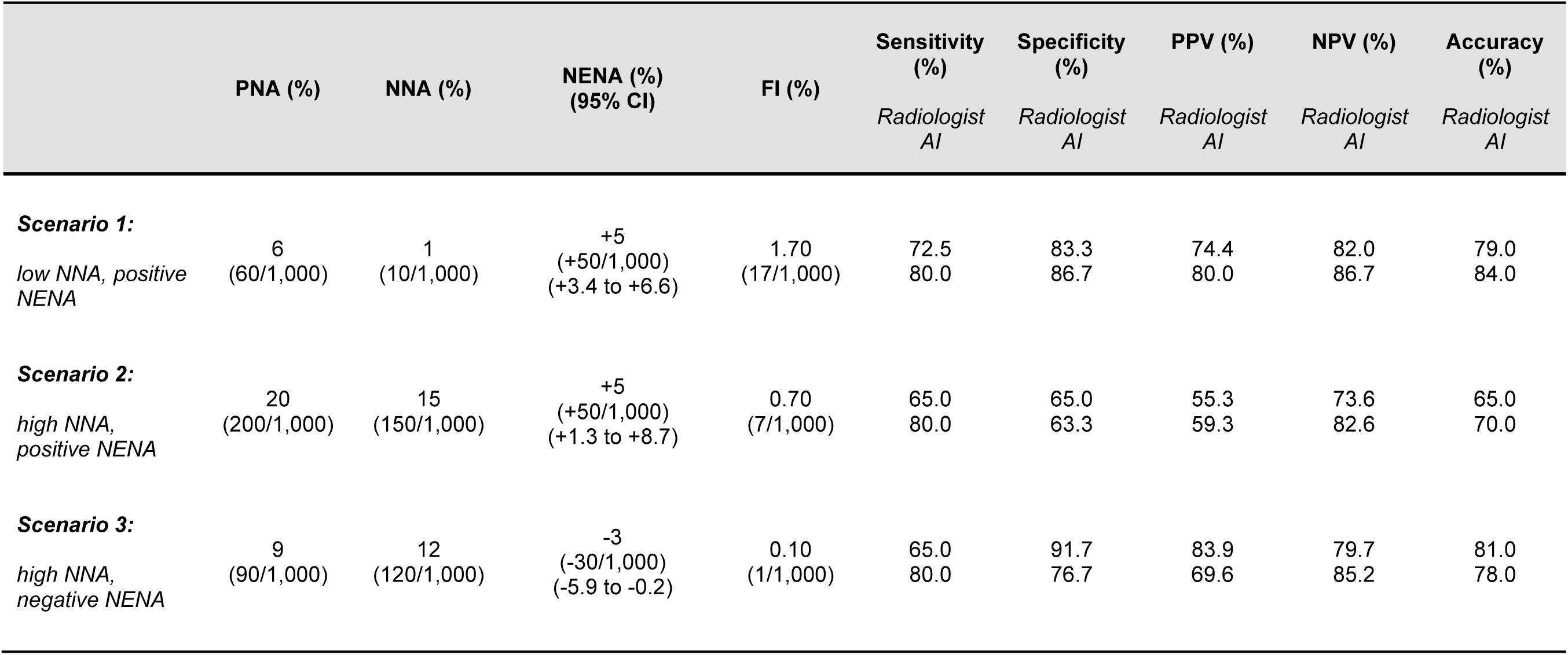
Hypothetical scenarios with directional reclassification metrics and conventional diagnostic performance metrics. The hypothetical scenarios use 1,000 examinations, including 400 reference-standard-positive and 600 reference-standard-negative cases. Conventional performance metrics are shown as AI *versus* radiologist values. Positive non-agreement (PNA), negative non-agreement (NNA), and net non-agreement (NENA) were calculated from paired AI-radiologist correctness values from the overall cohort. The 95% bootstrap confidence interval for NENA and the reclassification fragility index (FI) are also shown.

The 95% bootstrap confidence interval for NENA excluded 0 in all three scenarios, so the reclassification FI was defined for each (Table 2). Although scenarios 1 and 2 shared an identical NENA of +5%, the reclassification FI was 1.70% (17 out of 1,000 examinations) in scenario 1 (95% CI, +3.4% to +6.6%) and 0.70% (7 out of 1,000 examinations) in scenario 2 (95% CI, +1.3% to +8.7%), reflecting the wider confidence interval generated by the larger discordant counts of scenario 2 (PNA=200, NNA=150). In scenario 3 (NENA=-3%; 95% CI, −5.9% to −0.2%), reclassifying only one examination was sufficient for the confidence interval to include 0 (FI, 0.10%). With identical proportions but Q=100, the confidence interval for NENA included 0 in all three scenarios and the reclassification FI would not have been defined (Appendix A5).

### Applicability

Results of the application of the metric-triplet to the three selected studies are shown in Table 3, revealing that NENA varied with clinical setting as well as reference standard (5, 6). Directional non-agreement metrics also varied with reader’s experience (7). In the McKinney *et al.* cohort, NENA was positive for the first reader (+1.08%) but negative for the consensus decision (−4.18%), despite deriving from the same dataset (5). Across all seven comparisons, AI-platform sensitivity exceeded radiologist sensitivity, whereas NENA was negative in two comparisons (5–7). In the Goh *et al.* cohort, positive predictive value for the AI-platform was lower than for the radiologist across all three reader-experience groups, whereas NENA remained positive (7).

**Table 3.**
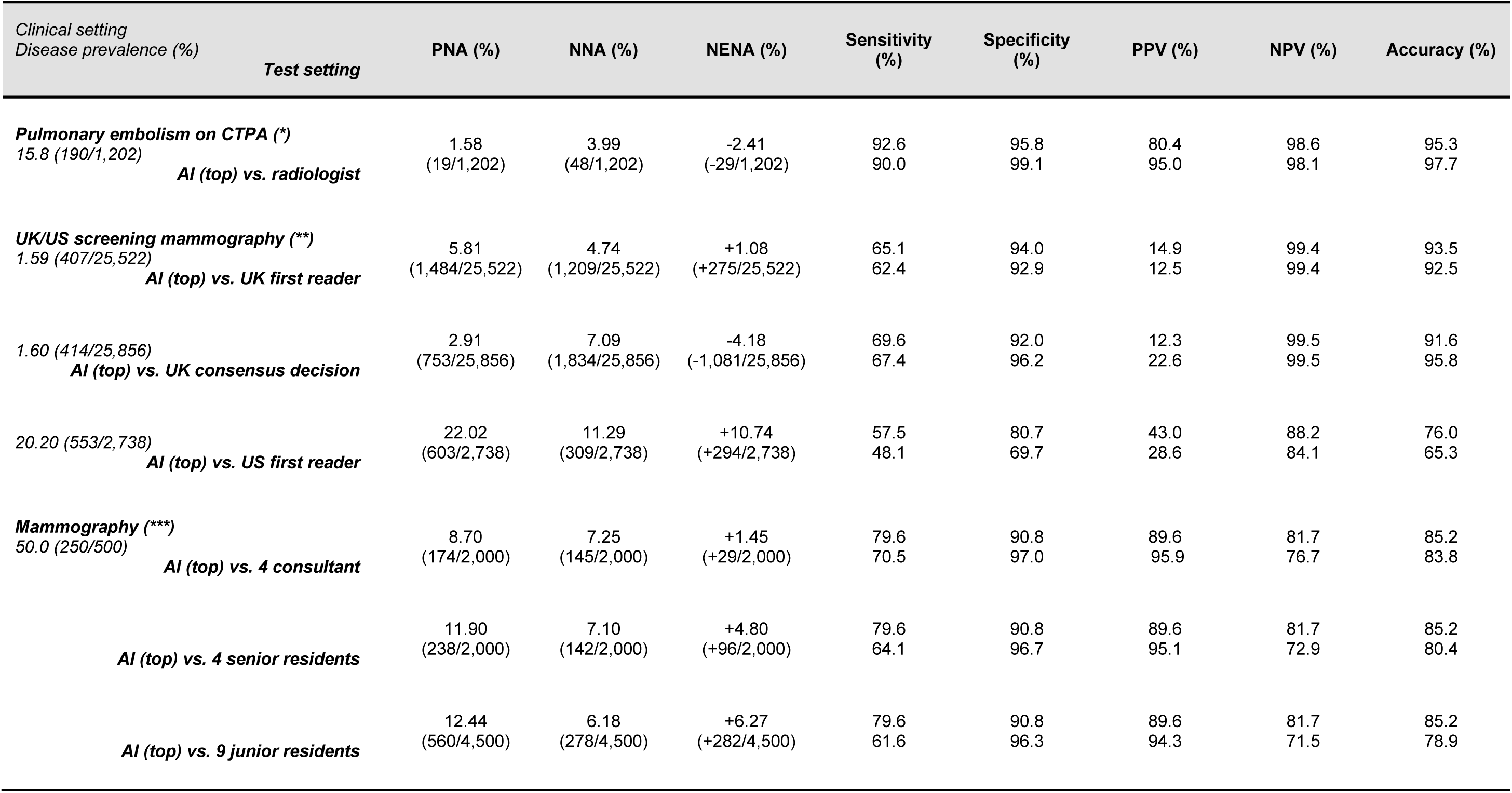
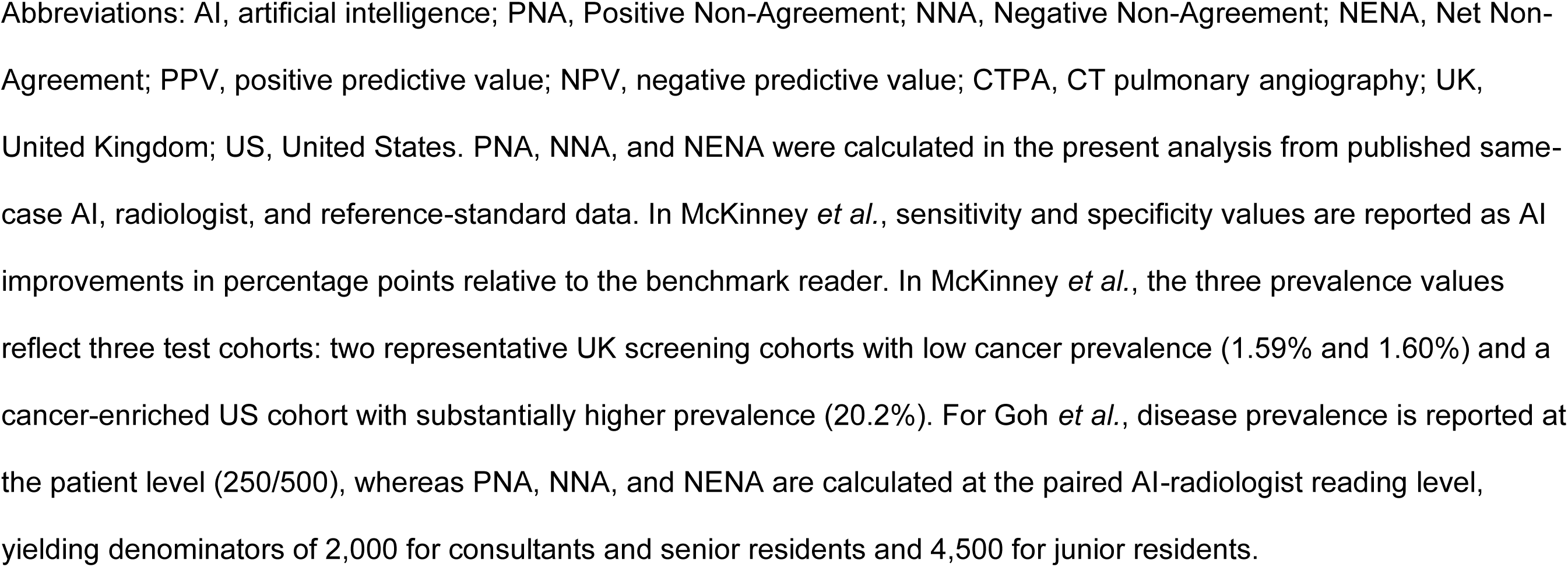
Directional metrics for applicability studies. Directional metrics are shown for all seven scenarios included in the three applicability studies, together with conventional performance metrics for comparison.

The reclassification FI was defined for six of the seven applicability comparisons and ranged from 0.34% (McKinney *et al.*, AI-platform *versus* first reader, low-prevalence cohort; 87 out of 25,522 examinations) to 4.31% (McKinney *et al.*, AI-platform *versus* first reader, high-prevalence cohort; 118 out of 2,738 examinations). The reclassification FI was not defined for comparisons between the AI-platform and consultant radiologists in the Goh *et al.* cohort, because its 95% bootstrap confidence interval for NENA (+1.45%; 95% CI, −0.35% to +3.20%) included 0. The two McKinney *et al.* comparisons with the largest cohorts (Q>25,000) had the smallest reclassification FI as a percentage of Q, but the largest in absolute examinations (87 and 491, respectively), reflecting greater statistical power in larger cohorts. Comparison-level reclassification FI values and a worked example are provided in Appendices A1 and A6.

## Discussion

In this study, we developed, illustrated, and applied a set of “directional” metrics to quantify non-agreements between radiologists and AI-platforms on the same individual examinations, as compared against a common reference standard. The resulting metric-triplet of PNA, NNA, and NENA identifies individual examinations in which the radiologist is right and the AI-platform is wrong, and *vice versa*, and quantifies the net effect of disagreement toward a beneficial or harmful effect of the AI-platform. We additionally adapted the Fragility Index, previously used to assess randomized trial robustness, into a reclassification metric that quantifies the minimum number of case reclassifications needed to remove statistical support for the direction of NENA. This set of “directional” metrics and the adapted reclassification FI complement ongoing efforts to make AI-platform outputs more interpretable and trustworthy for radiologists (18).

Performance metrics, such as sensitivity, specificity, accuracy, and predictive values quantify standalone radiologist or AI-platform performance against an independent reference standard (19). Kappa statistics quantify how often readers and/or methods disagree beyond chance, without comparison against any reference standard, so whether that disagreement reflects being right or wrong remains unknown (20–22). PNA and NNA identify which non-agreement examinations are driven by a beneficial or harmful AI-platform decision, with NENA summarizing their net balance. Because two comparisons can share similar overall accuracy while differing in PNA, NNA, and NENA, a radiology department may need these figures to determine whether a favorable accuracy conceals a non-trivial rate of AI-introduced errors. NENA should always be interpreted together with PNA and NNA, since a positive NENA alone does not guarantee that harmful disagreements are acceptably rare.

PNA and NNA are the normalized frequencies of both discordant cells of a paired comparison, the same cells that McNemar’s test compares in paired AI-platform-radiologist accuracy studies (13). NENA is correspondingly identical to the paired difference in accuracy. Our contribution is, therefore, not a new statistical test but a reframing of existing tests for a specific purpose. First, PNA and NNA are labeled by clinical direction, beneficial or harmful, rather than an undifferentiated discordance count. Indeed, a McNemar p-value indicates whether an AI-platform and a radiologist differ, but collapses both directions into a single statistic, unable to distinguish a beneficial-dominant pattern from a harmful-dominant one. Second, decomposing accuracy differences into PNA and NNA identifies why an AI-platform’s accuracy differs from a radiologist’s accuracy, not only whether it differs, as illustrated by scenarios 1 and 2. These scenarios share an identical NENA but differ markedly in NNA. Third, we position PNA, NNA, and NENA as an AI evaluation and deployment-monitoring tool rather than as a hypothesis test between two classifiers. Discordant-pair testing establishes whether an AI-platform and a radiologist differ, while our framework additionally specifies the direction of that difference and its likely clinical consequence. Fourth, the reclassification FI extends this framework beyond relabeling. By quantifying the minimum number of case reclassifications needed to remove statistical support for the direction of a metric, evaluated through its bootstrap confidence interval rather than its point estimate, it has no direct precedent in the McNemar literature, and could apply to any paired binary comparison against a reference standard.

The metric-triplet accompanied by the reclassification FI could have implications for deployment decisions. Consider two departments that each observe an AI-platform accuracy 5 percentage points higher than their radiologists, corresponding to the identical NENA of +5% shared by scenarios 1 and 2. In the first department (scenario 1, NNA 1%), 10 out of 1,000 examinations would be misclassified because of AI-platform error, and unsupervised use of AI may have limited risk. In the second department (scenario 2, NNA 15%), 150 out of 1,000 examinations would be misclassified. Reporting NENA alone would treat these situations as equivalent, whereas reporting PNA and NNA shows the second department requires a more cautious approach despite an identical net accuracy gain. The reclassification FI adds a further distinction: with 1,000 examinations per scenario, the positive NENA of scenario 1 would lose statistical support only after 17 examinations were reclassified, whereas that of scenario 2 would do so after 7. The same occurred in real data. In the study from Ben Cheikh *et al*., the AI-platform had a higher sensitivity than the radiologist and a similar negative predictive value, yet NENA was negative because NNA (radiologist right) outnumbered PNA (radiologist wrong), a finding not captured by sensitivity or negative predictive value alone and likely cautioning against AI deployment given the higher potential for harmful non-agreements (6). The reclassification FI for this comparison was 0.58% (seven out of 1,202 examinations), the second lowest of the six defined values in our applicability series. Reclassifying seven examinations would remove statistical support for this conclusion, which is particularly notable since Ben Cheikh *et al*. also had the least independent reference standard among the three applicability studies.

The reclassification FI results have several implications beyond the specific study by Ben Cheikh *et al*. First, anchoring the reclassification FI to the 95% CI for NENA rather than the NENA point estimate is what allows it to convey information beyond NENA itself (Appendix A1). A point-estimate-based count cannot distinguish a well-powered from a poorly powered conclusion with the same point estimate, whereas our proposed definition can, as shown by our results. The McKinney *et al*. comparison against the consensus decision has a smaller effect (NENA=-4.18%) than against the first reader in the higher-prevalence cohort (NENA=+10.74%), yet its larger sample (Q=25,856 *versus* Q=2,738) requires reclassifying more cases in absolute terms to lose statistical support (491 *versus* 118), despite a smaller reclassification FI as a percentage of Q (1.90% *versus* 4.31%) (5). Conversely, for a fixed NENA, the reclassification FI expressed as a percentage of Q increases with sample size toward approximately half the absolute value of NENA, the value of the point-estimate-based count (Appendix A5). Second, the reclassification FI was undefined for the Goh *et al*. comparison against consultant radiologists because its confidence interval for NENA included zero (7), whereas it was defined for all three illustrative scenarios only because of their cohort size: with identical proportions but 100 instead of 1,000 examinations, none of their confidence intervals for NENA would have excluded zero (Appendix A5). The reclassification FI thus retains a dual interpretation, reflecting both reference-standard tolerance and statistical power, arguing against overreliance on conclusions from either an imperfect reference standard or an underpowered comparison.

Other observations from our study have similar implications for implementing a given AI-platform. In the study by Goh *et al*., NENA continuously decreased from junior residents (+6.27%) to senior residents (+4.80%) and consultant radiologists (+1.45%), inversely proportional to their experience, paralleling findings that the incremental benefit of second readers, either human or AI-based, tends to be greatest for less experienced readers (7, 23, 24). This can be of interest in departments with high numbers of radiologists in training, whereas in departments without trainees the appeal of AI implementation might be marginal. In addition to reader’s experience, the metric-triplet could also stratify by examination type, case complexity, or workflow demands, complementing sensitivity, specificity, and accuracy in deployment decisions (12, 25). Because PNA and NNA translate directly into counts of examinations correctly and incorrectly reclassified by an AI-platform, our observations could also inform estimations of return on investment of AI-platform implementation, potentially weighing these counts against the costs of missed diagnoses or unnecessary follow-up examinations.

The reference standard is of pivotal importance, and its variability could have influenced the directional metrics. In the study by Ben Cheikh *et al.*, the final diagnosis was determined by retrospective expert review of the CT examinations, informed by clinical information, radiology reports, AI results, and patient outcomes, so the reference standard was co-informed by both AI and radiologist interpretations and not completely independent (6). In McKinney *et al*. and Goh *et al*., positive cases required a biopsy-confirmed diagnosis and negative cases a cancer-free screening examination or benign biopsy at up to 24 months of follow-up, a reference standard likely less directly influenced by radiologist and AI assessments than in the study by Ben Cheikh *et al.*, although clinical interpretations could still have contributed to the decision to biopsy (5–7). This variability in reference-standard definition is a recognized limitation across AI validation studies (26). These differences also partly explain the variability of the metric-triplet across studies, unrelated to the AI-platform itself. Since its accuracy depends on the independence and robustness of the reference standard, PNA, NNA, and NENA should always be reported together with its description, without which these values cannot be appropriately interpreted.

The proposed metric-triplet has several limitations. First, the metric-triplet depends on an independent and robust reference standard. Without it, any given non-agreement can reflect a true miss by either the radiologist or the AI-platform, or an attribution error of the reference standard itself. This calls for caution when the reference standard is retrospective, poorly validated, or not fully independent from either assessment (27, 28). The reclassification FI provides a complementary assessment of the robustness of directional conclusions to case reclassification but does not eliminate reference-standard bias. Second, the metric-triplet quantifies AI-platforms’ potential to either decrease or increase radiologists’ reading errors, but its clinical significance depends on whether radiologists notice, accept, or reject AI-platform output, and on whether disagreement changes management (29–31). Automation bias could amplify this effect if not adequately monitored (11, 32), an aspect not captured by our study design. Third, the metric-triplet is currently limited to binary outcomes. Continuous measurements or scores would require prespecified thresholds or categorization before PNA, NNA, and NENA could be calculated. Finally, the metric-triplet weights all non-agreement equally and does not account for clinical relevance, including management decisions, workflow overreliance, and model drift (30, 31, 33). A missed pulmonary embolism and an unnecessary mammography recall are both treated as a single PNA event despite differing urgency and resource consequences, and the same applies to NNA, where a missed cancer misattributed to an AI-platform error carries greater risk than an overridden false-positive. A future iteration of our approach could weigh PNA and NNA by clinical cost, via decision curve analysis or multimodal foundation-model outputs (34, 35).

In conclusion, the metric-triplet consisting of PNA, NNA, and NENA provides a directional approach for quantifying non-agreement between radiologists and AI-platforms, and for determining whether this non-agreement tends to be beneficial or harmful in each clinical setting. These metrics complement conventional performance metrics and provide radiology departments, vendors, and regulators with an easy-to-use and setting-specific tool for AI-evaluation, deployment decisions, and post-deployment monitoring. The accompanying reclassification FI is an additional tool for quantifying the robustness of directional conclusions and may find use beyond the metric-triplet for other paired diagnostic comparisons.

## Code Availability

The notation and formulas are provided in Appendix A1, and the R code is provided in Appendix A7.

## Funding

This study received no funding.

## Competing Interests

The authors declare no competing interests.

## Ethics Statement

This study analyzed hypothetical scenarios and aggregate summary data of previously published peer-reviewed articles. No human participants were enrolled and no individual-level data were accessed; institutional review board approval and informed consent were therefore not required.

## Abbreviations

AI: artificial intelligence;
PNA: Positive Non-Agreement;
NNA: Negative Non-Agreement;
NENA: Net Non-Agreement;
FI: fragility index;
CI: confidence interval;
CTPA: CT pulmonary angiography;
PPV: positive predictive value;
NPV: negative predictive value.

## Data Availability

All data produced in the present study are available upon reasonable request to the authors.

This study did not generate new patient-level data. All PNA, NNA, and NENA values applied to published data were calculated from previously reported, publicly available summary statistics extracted from the cited studies (5–7), as detailed in Table 3.

---

Figure 1. Figure shows Sankey diagrams of the illustrative scenarios 1 to 3 (Figure 1A to 1C), summarizing the absolute numbers changing between radiologist and AI-platform in terms of true-positive (TP), false-negative (FN), false-positive (FP), and true-negative (TN) assessments. Related sensitivity, specificity, accuracy, and predictive values are given in complementing Table 2.

**Figure 1A.**
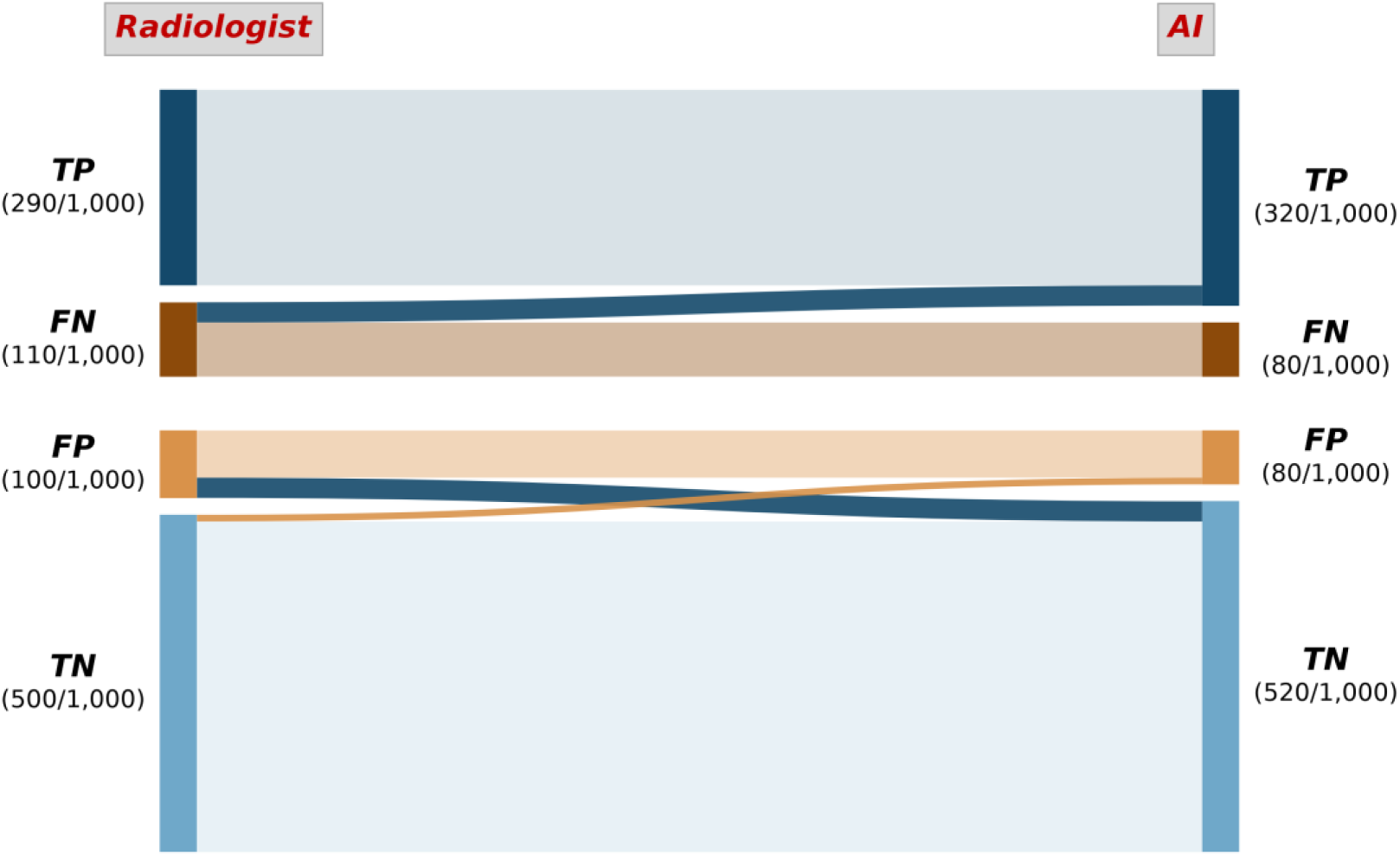
Scenario 1. Sankey diagram of the first scenario, with fractions of absolute numbers. In this scenario, PNA was 60/1,000 (6%), NNA was 10/1,000 (1%), and NENA was +50/1,000 (+5%).

**Figure 1B.**
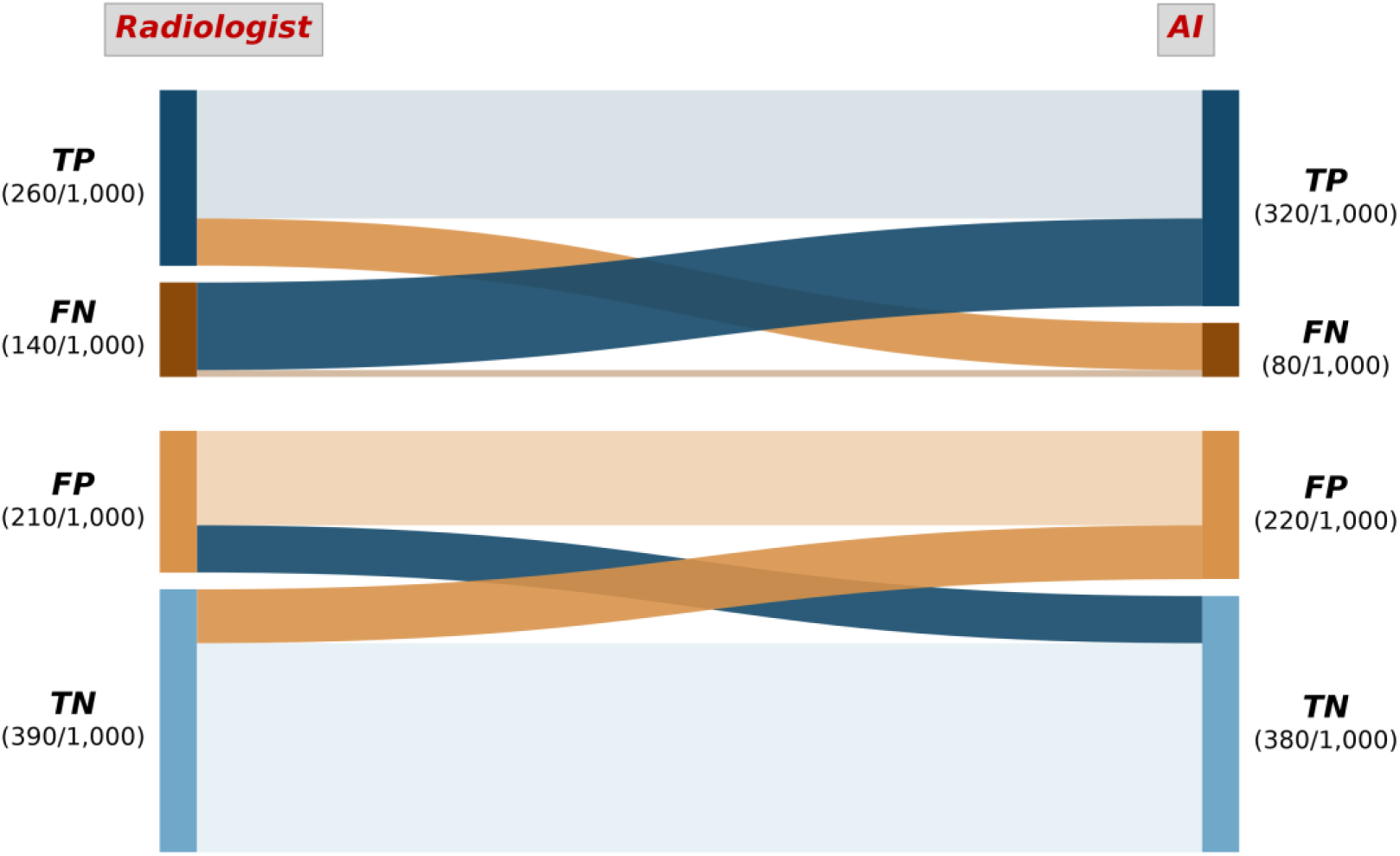
Scenario 2. Sankey diagram of the second scenario, with fractions of absolute numbers. In this scenario, PNA was 200/1,000 (20%), NNA was 150/1,000 (15%), and NENA was +50/1,000 (5%).

**Figure 1C.**
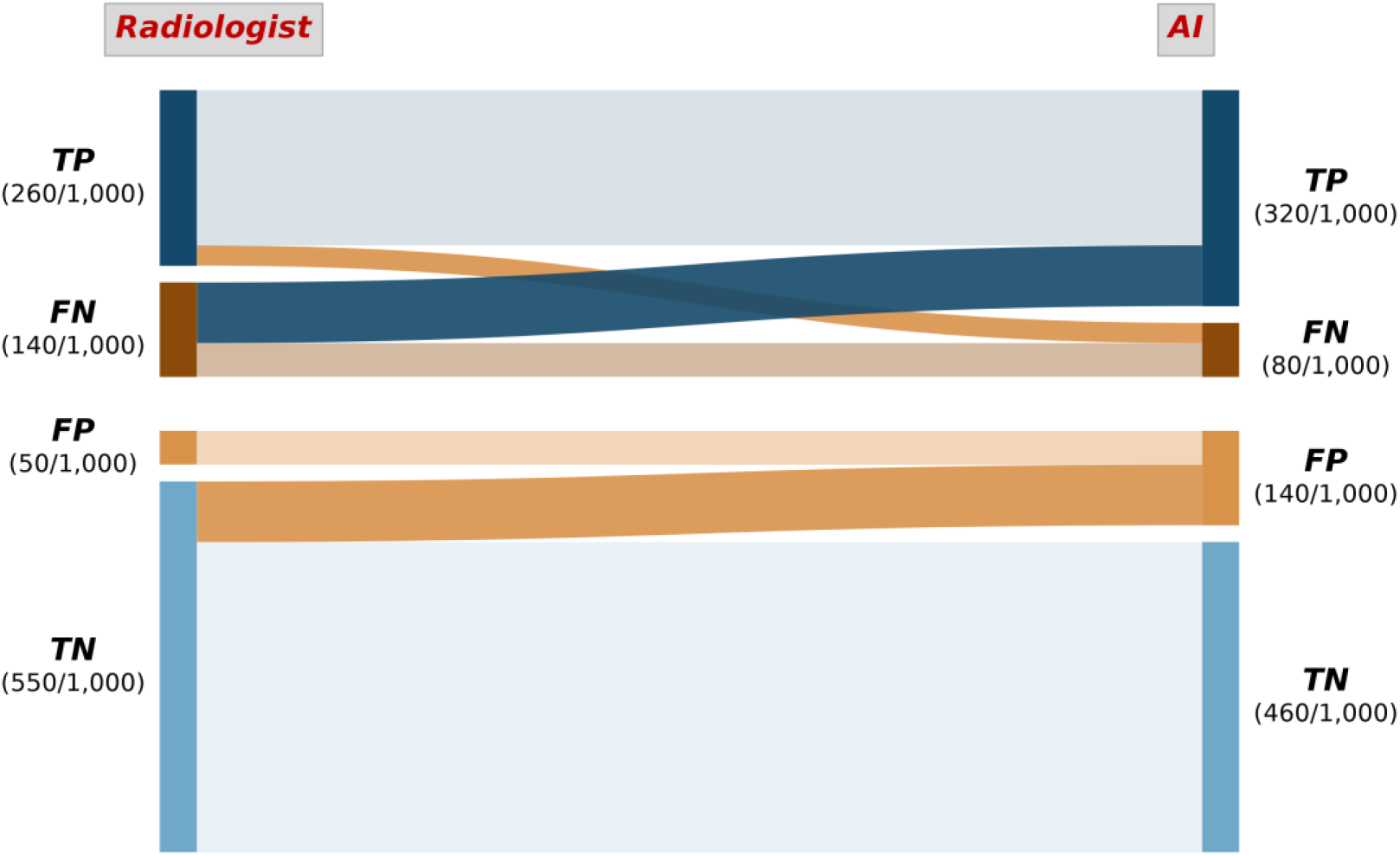
Scenario 3. Sankey diagram of the third scenario, with fractions of absolute numbers. In this scenario, PNA was 90/1,000 (9%), NNA was 120/1,000 (12%), and NENA was −30/1,000 (−3%).

## Notes

### Competing Interest Statement

The authors have declared no competing interest.

## References

1. Topol EJ. High-performance medicine: the convergence of human and artificial intelligence. Nat Med 2019;25(1):44–56. doi: 10.1038/s41591-018-0300-7

2. Trevethan R. Sensitivity, Specificity, and Predictive Values: Foundations, Pliabilities, and Pitfalls in Research and Practice. Front Public Health 2017;5:307. doi: 10.3389/fpubh.2017.00307

3. Rodriguez-Ruiz A, Lang K, Gubern-Merida A, Broeders M, Gennaro G, Clauser P, Helbich TH, Chevalier M, Tan T, Mertelmeier T, Wallis MG, Andersson I, Zackrisson S, Mann RM, Sechopoulos I. Stand-Alone Artificial Intelligence for Breast Cancer Detection in Mammography: Comparison With 101 Radiologists. J Natl Cancer Inst 2019;111(9):916–922. doi: 10.1093/jnci/djy222

4. Aggarwal R, Sounderajah V, Martin G, Ting DSW, Karthikesalingam A, King D, Ashrafian H, Darzi A. Diagnostic accuracy of deep learning in medical imaging: a systematic review and meta-analysis. NPJ Digit Med 2021;4(1):65. doi: 10.1038/s41746-021-00438-z

5. McKinney SM, Sieniek M, Godbole V, Godwin J, Antropova N, Ashrafian H, Back T, Chesus M, Corrado GS, Darzi A, Etemadi M, Garcia-Vicente F, Gilbert FJ, Halling-Brown M, Hassabis D, Jansen S, Karthikesalingam A, Kelly CJ, King D, Ledsam JR, Melnick D, Mostofi H, Peng L, Reicher JJ, Romera-Paredes B, Sidebottom R, Suleyman M, Tse D, Young KC, De Fauw J, Shetty S. International evaluation of an AI system for breast cancer screening. Nature 2020;577(7788):89–94. doi: 10.1038/s41586-019-1799-6

6. Ben Cheikh A, Gorincour G, Nivet H, May J, Seux M, Calame P, Thomson V, Delabrousse E, Crombe A. How artificial intelligence improves radiological interpretation in suspected pulmonary embolism. Eur Radiol 2022;32(9):5831–5842. doi: 10.1007/s00330-022-08645-2

7. Goh SSN, Du H, Tan LY, Seah EZY, Lau WK, Ng AHZ, Lim SWD, Ong HY, Lau S, Tan YL, Khaw MS, Yap CW, Hui KYD, Tan WC, Abdul H, Khoo VMH, Ge S, Pool FJ, Choo YS, Wang Y, Jagmohan P, Gopinathan PP, Hartman M, Feng M. Impact of AI on Breast Cancer Detection Rates in Mammography by Radiologists of Varying Experience Levels in Singapore: Preliminary Comparative Study. JMIR Form Res 2025;9:e66931. doi: 10.2196/66931

8. Muehlematter UJ, Daniore P, Vokinger KN. Approval of artificial intelligence and machine learning-based medical devices in the USA and Europe (2015-20): a comparative analysis. Lancet Digit Health 2021;3(3):e195–e203. doi: 10.1016/S2589-7500(20)30292-2

9. Kelly CJ, Karthikesalingam A, Suleyman M, Corrado G, King D. Key challenges for delivering clinical impact with artificial intelligence. BMC Med 2019;17(1):195. doi: 10.1186/s12916-019-1426-2

10. Recht MP, Dewey M, Dreyer K, Langlotz C, Niessen W, Prainsack B, Smith JJ. Integrating artificial intelligence into the clinical practice of radiology: challenges and recommendations. Eur Radiol 2020;30(6):3576–3584. doi: 10.1007/s00330-020-06672-5

11. Lyell D, Coiera E. Automation bias and verification complexity: a systematic review. J Am Med Inform Assoc 2017;24(2):423–431. doi: 10.1093/jamia/ocw105

12. Pesapane F, Latronico A, Abbate F, Penco S, Rotili A, Dominelli V, Nicosia L, Monzani D, Grasso R, Pravettoni G, Cassano E. Evaluating cognitive biases in AI-assisted mammography interpretation: a simulation reader study of explainable AI across radiologist experience levels. Eur Radiol 2026;36(10):7639–7650. doi: 10.1007/s00330-026-12666-6

13. Mc Nemar Q. Note on the sampling error of the difference between correlated proportions or percentages. Psychometrika 1947;12(2):153–157. doi: 10.1007/BF02295996

14. Efron B. Bootstrap Methods: Another Look at the Jackknife. The Annals of Statistics 1979;7(1). doi: 10.1214/aos/1176344552

15. Efron B, Tibshirani RJ. An Introduction to the Bootstrap. New York: Chapman & Hall, 1993.

16. Field CA, Welsh AH. Bootstrapping Clustered Data. Journal of the Royal Statistical Society Series B: Statistical Methodology 2007;69(3):369–390. doi: 10.1111/j.1467-9868.2007.00593.x

17. Walsh M, Srinathan SK, McAuley DF, Mrkobrada M, Levine O, Ribic C, Molnar AO, Dattani ND, Burke A, Guyatt G, Thabane L, Walter SD, Pogue J, Devereaux PJ. The statistical significance of randomized controlled trial results is frequently fragile: a case for a Fragility Index. J Clin Epidemiol 2014;67(6):622–628. doi: 10.1016/j.jclinepi.2013.10.019

18. Reyes M, Meier R, Pereira S, Silva CA, Dahlweid FM, von Tengg-Kobligk H, Summers RM, Wiest R. On the Interpretability of Artificial Intelligence in Radiology: Challenges and Opportunities. Radiol Artif Intell 2020;2(3):e190043. doi: 10.1148/ryai.2020190043

19. Obuchowski NA, Bullen JA. Receiver operating characteristic (ROC) curves: review of methods with applications in diagnostic medicine. Phys Med Biol 2018;63(7):07TR01. doi: 10.1088/1361-6560/aab4b1

20. Cohen J. A Coefficient of Agreement for Nominal Scales. Educational and Psychological Measurement 1960;20(1):37–46. doi: 10.1177/001316446002000104

21. Landis JR, Koch GG. The Measurement of Observer Agreement for Categorical Data. Biometrics 1977;33(1). doi: 10.2307/2529310

22. Kundel HL, Polansky M. Measurement of observer agreement. Radiology 2003;228(2):303–308. doi: 10.1148/radiol.2282011860

23. Lang K, Josefsson V, Larsson AM, Larsson S, Hogberg C, Sartor H, Hofvind S, Andersson I, Rosso A. Artificial intelligence-supported screen reading versus standard double reading in the Mammography Screening with Artificial Intelligence trial (MASAI): a clinical safety analysis of a randomised, controlled, non-inferiority, single-blinded, screening accuracy study. Lancet Oncol 2023;24(8):936–944. doi: 10.1016/S1470-2045(23)00298-X

24. Taylor-Phillips S, Stinton C. Double reading in breast cancer screening: considerations for policy-making. Br J Radiol 2020;93(1106):20190610. doi: 10.1259/bjr.20190610

25. Dembrower K, Wahlin E, Liu Y, Salim M, Smith K, Lindholm P, Eklund M, Strand F. Effect of artificial intelligence-based triaging of breast cancer screening mammograms on cancer detection and radiologist workload: a retrospective simulation study. Lancet Digit Health 2020;2(9):e468–e474. doi: 10.1016/S2589-7500(20)30185-0

26. Freeman K, Geppert J, Stinton C, Todkill D, Johnson S, Clarke A, Taylor-Phillips S. Use of artificial intelligence for image analysis in breast cancer screening programmes: systematic review of test accuracy. BMJ 2021;374:n1872. doi: 10.1136/bmj.n1872

27. Elmore JG, Wells CK, Lee CH, Howard DH, Feinstein AR. Variability in radiologists’ interpretations of mammograms. N Engl J Med 1994;331(22):1493–1499. doi: 10.1056/NEJM199412013312206

28. Bruno MA, Walker EA, Abujudeh HH. Understanding and Confronting Our Mistakes: The Epidemiology of Error in Radiology and Strategies for Error Reduction. Radiographics 2015;35(6):1668–1676. doi: 10.1148/rg.2015150023

29. Zech JR, Badgeley MA, Liu M, Costa AB, Titano JJ, Oermann EK. Variable generalization performance of a deep learning model to detect pneumonia in chest radiographs: A cross-sectional study. PLoS Med 2018;15(11):e1002683. doi: 10.1371/journal.pmed.1002683

30. Goddard K, Roudsari A, Wyatt JC. Automation bias: a systematic review of frequency, effect mediators, and mitigators. Journal of the American Medical Informatics Association 2012;19(1):121–127. doi: 10.1136/amiajnl-2011-000089

31. Gaube S, Suresh H, Raue M, Merritt A, Berkowitz SJ, Lermer E, Coughlin JF, Guttag JV, Colak E, Ghassemi M. Do as AI say: susceptibility in deployment of clinical decision-aids. NPJ Digit Med 2021;4(1):31. doi: 10.1038/s41746-021-00385-9

32. Cabitza F, Rasoini R, Gensini GF. Unintended Consequences of Machine Learning in Medicine. JAMA 2017;318(6):517–518. doi: 10.1001/jama.2017.7797

33. Vokinger KN, Feuerriegel S, Kesselheim AS. Continual learning in medical devices: FDA’s action plan and beyond. Lancet Digit Health 2021;3(6):e337–e338. doi: 10.1016/S2589-7500(21)00076-5

34. Vickers AJ, Elkin EB. Decision curve analysis: a novel method for evaluating prediction models. Med Decis Making 2006;26(6):565–574. doi: 10.1177/0272989X06295361

35. Moor M, Banerjee O, Abad ZSH, Krumholz HM, Leskovec J, Topol EJ, Rajpurkar P. Foundation models for generalist medical artificial intelligence. Nature 2023;616(7956):259–265. doi: 10.1038/s41586-023-05881-4

