## Supplementary Appendices A1-A7 for "Disagreement between AI and Radiologists: Quantifying Direction and Fragility"

### APPENDIX A1

#### Reclassification Fragility Index: derivation, procedure, and example

Notation. In all appendices, Q denotes the total number of paired assessments (examinations or readings) in a comparison, P the number of assessments in which the AI-platform was right and the radiologist wrong (positive non-agreement events), and N the number of assessments in which the radiologist was right and the AI-platform wrong (negative non-agreement events), both as compared against the reference standard. Positive Non-Agreement (PNA)=P/Q, Negative Non-Agreement (NNA)=N/Q, and Net Non-Agreement (NENA)=PNA-NNA=(P-N)/Q. TP, FN, FP, and TN denote true-positive, false-negative, false-positive, and true-negative classifications, respectively; PPV and NPV denote positive and negative predictive values; CI denotes confidence interval; CTPA denotes CT pulmonary angiography; and UK and US denote the United Kingdom and the United States.

A Fragility Index (FI), defined as the number of case reclassifications needed to move the point estimate of NENA to 0, is fully determined by P and N. Each case reclassified from the majority discordant category to the minority category changes P-N by 2. Therefore, a point-estimate-based fragility count is directly determined by the size of NENA and adds no independent information. To provide information beyond the point estimate, we instead anchored the reclassification FI to the 95% bootstrap confidence interval for NENA.

Each assessment contributing to Q was modeled as belonging to one of three categories: positive non-agreement (PNA, probability P/Q); negative non-agreement

(NNA, probability N/Q); or concordant (both radiologist and AI-platform right, or both wrong, probability 1-P/Q-N/Q). Because the concordant category is not subdivided, this bootstrap procedure requires only the counts P, N, and Q, and not case-level data or knowledge of how concordant examinations divide between both-right and both-wrong (34). In multinomial resampling, the joint distribution of any subset of categories depends only on those categories' probabilities and the total sample size, regardless of how the remaining probability is partitioned (34). This is what allowed the reclassification FI to be calculated identically for the illustrative scenarios and for the applicability studies, the latter extracted from aggregate published data without access to case-level records.

For each comparison, Q assessments were resampled with replacement 10,000 times according to these category probabilities. NENA was recalculated for each resample as (P′-N′)/Q, and the 2.5th and 97.5th percentiles of the resulting distribution formed the 95% confidence interval (34). If this baseline interval included 0, the reclassification FI was considered not applicable. If it excluded 0, cases were reclassified one at a time from the majority discordant category to the minority category. The bootstrap confidence interval was recalculated at each step using the modified counts, and the process continued until the recalculated interval first included 0. The number of reclassified cases at that point, expressed as a percentage of Q, is the reclassification FI. Across every comparison evaluated, the position of the confidence interval relative to 0 changed monotonically as the number of reclassified cases increased, so a binary search over the number of reclassified cases was used instead of an exhaustive linear search. An implementation requires only a routine capable of multinomial random sampling and percentile-based interval calculation, available in standard statistical software including R.

This procedure treated each assessment contributing to Q as an independent observation. If assessments were correlated, for example when several radiologists evaluated the same examination, the effective sample size might be smaller than Q. This might affect the bootstrap confidence interval and the reclassification FI. Because the published applicability studies provided only aggregate data, we could not adjust for this clustering.

For example, in the comparison from Ben Cheikh *et al.* (P=19, N=48, Q=1,202, NENA=-2.41%), the baseline 95% bootstrap confidence interval for NENA was -3.74% to -1.08%, which excluded 0. Reclassifying cases from NNA to PNA, the majority-to-minority direction appropriate for a negative NENA, moved N down and P up by one case at a time. Recalculating the bootstrap confidence interval after each reclassification showed the interval narrowing and shifting toward 0. After 7 reclassifications (P=26, N=41), the recalculated 95% confidence interval first included 0, so the reclassification FI for this comparison was 7/1,202 = 0.58%. This means that if as few as 7/1,202 assessments in this comparison had been reclassified in the opposite direction, plausibly through reference-standard error, given that Ben Cheikh *et al*. used a reference standard partially informed by AI and radiologist interpretations, the directional conclusion that the AI-platform introduced more harmful than beneficial non-agreement would no longer be supported.

### APPENDIX A2

#### Reconstruction of data from Ben Cheikh *et al*.

*Ben Cheikh et al.: pulmonary embolism on CT pulmonary angiography*

Ben Cheikh *et al*. evaluated 1,202 CTPA examinations. According to Table 1 of the original article, 190 examinations were positive for pulmonary embolism (PE) as compared against the reference standard, corresponding to a disease prevalence of 15.8% (190/1,202). The remaining 1,012 examinations were therefore reference-standard negative.

This article provided sufficient aggregate and same-case information to reconstruct the conventional diagnostic performance of the AI algorithm and the radiologists as well as the direction of their disagreement relative to the reference standard.

The authors reported that the AI algorithm identified 219 examinations as suspicious for PE, including 176 TP and 43 FP examinations. Fourteen reference-standard-positive examinations were missed by AI and were therefore false negatives (FN). Because 1,012 examinations were reference-standard negative, the number of AI TN examinations was reconstructed as:

$$TN_{AI}=1{,}012-43=969$$

The resulting reference-standard-by-AI confusion matrix was:

| Reference standard | AI positive | AI negative | Total |
|---|---|---|---|
| PE present | 176 (TP) | 14 (FN) | 190 |
| PE absent | 43 (FP) | 969 (TN) | 1,012 |
| Total | 219 | 983 | 1,202 |

For the radiologists, the authors reported 180 examinations considered suspicious for PE, including 171 TP and 9 FP examinations. Nineteen reference-standard-positive examinations were missed by the radiologists and were therefore FN. The number of radiologist TN examinations was consequently reconstructed as:

$$TN_{Rad}=1{,}012-9=1{,}003$$

The resulting reference-standard-by-radiologist confusion matrix was:

| Reference standard | Radiologist positive | Radiologist negative | Total |
|---|---|---|---|
| PE present | 171 (TP) | 19 (FN) | 190 |
| PE absent | 9 (FP) | 1,003 (TN) | 1,012 |
| Total | 180 | 1,022 | 1,202 |

These confusion matrices establish the standalone performance of AI and the radiologists but do not, by themselves, identify whether their errors occurred in the same examinations. The article additionally provided sufficient same-case information to reconstruct the paired AI-radiologist matrices separately for reference-standard-positive and reference-standard-negative examinations.

*Reconstruction among reference-standard-positive examinations*

Among the 190 examinations with PE, the authors reported that 19 true PEs missed by the radiologists were detected by AI. These examinations therefore corresponded to AI TP/radiologist FN discordance. Conversely, 14 true PEs missed by AI were detected by the radiologists, corresponding to AI FN/radiologist TP discordance.

AI had 176 TP examinations in total. After subtracting the 19 examinations in which AI was TP and the radiologists were FN, 157 examinations remained in which both AI and the radiologists were TP:

$$176-19=157$$

The same value was obtained from the radiologists' totals:

$$171-14=157$$

The paired matrix for the 190 reference-standard-positive examinations was therefore reconstructed as:

| | Radiologist TP | Radiologist FN | Total |
|---|---|---|---|
| AI TP | 157 | 19 | 176 |
| AI FN | 14 | 0 | 14 |
| Total | 171 | 19 | 190 |

Thus, among reference-standard-positive examinations, 19 examinations represented PNA because AI was correct while the radiologists were incorrect. Fourteen examinations represented NNA because the radiologists were correct while AI was incorrect.

*Reconstruction among reference-standard-negative examinations*

Among the 1,012 reference-standard-negative examinations, the authors further reported that AI falsely indicated PE in 34 patients in whom the radiologists correctly indicated the absence of PE. These 34 examinations therefore corresponded to AI FP/radiologist TN discordance.

AI had 43 FP examinations in total. Subtracting the 34 examinations in which AI was FP while the radiologists were TN left 9 examinations in which both AI and the radiologists were FP:

$$43-34=9$$

Because the radiologists had 9 FP examinations in total, all radiologist FP examinations were also classified as FP by AI. Consequently, there were no examinations in which AI was TN while the radiologists were FP.

The paired matrix for the 1,012 reference-standard-negative examinations was therefore reconstructed as:

| | Radiologist TN | Radiologist FP | Total |
|---|---|---|---|
| AI TN | 969 | 0 | 969 |
| AI FP | 34 | 9 | 43 |
| Total | 1,003 | 9 | 1,012 |

Among reference-standard-negative examinations, no additional PNA events occurred. The 34 AI FP/radiologist TN examinations represented additional NNA events because the radiologists were correct while AI was incorrect.

*Calculation of PNA, NNA, and NENA*

In the present analysis, PNA was defined as the proportion of examinations for which AI was correct and the radiologists were incorrect, whereas NNA was defined as the proportion for which the radiologists were correct and AI was incorrect. NENA was calculated as the difference between PNA and NNA:

$$PNA=P/Q$$

$$NNA=N/Q$$

$$NENA=(P-N)/Q$$

These definitions correspond to the directional non-agreement framework used throughout the study.

For Ben Cheikh *et al.*, the only PNA events were the 19 reference-standard-positive examinations in which AI detected PE and the radiologists missed it:

$$\mathrm{PNA}=\frac{19}{1{,}202}=1.58\%$$

NNA consisted of two components: 14 reference-standard-positive examinations in which the radiologists detected PE and AI missed it, and 34 reference-standard-negative examinations in which the radiologists correctly excluded PE but AI falsely indicated it:

$$\mathrm{NNA}=\frac{14+34}{1{,}202}=\frac{48}{1{,}202}=3.99\%$$

NENA was therefore:

$$\begin{aligned}&\mathrm{NENA}=\mathrm{PNA}-\mathrm{NNA}\\&=\frac{19-48}{1{,}202}\\&=\frac{-29}{1{,}202}=-2.41\%\end{aligned}$$

Thus, AI-radiologist disagreements favored the radiologists overall in this cohort: AI correctly reclassified 19 examinations that the radiologists classified incorrectly, whereas the radiologists correctly classified 48 examinations that AI classified incorrectly.

*Reconstruction and verification of conventional performance metrics*

The reconstructed marginal confusion matrices were also used to independently calculate the sensitivity, specificity, PPV, NPV, and accuracy for both AI and radiologists.

For AI:

$$\text{Sensitivity}_{\text{AI}}=\frac{176}{176+14}=\frac{176}{190}=92.6\%$$

$$\text{Specificity}_{\text{AI}}=\frac{969}{969+43}=\frac{969}{1{,}012}=95.8\%$$

$$\text{PPV}_{\text{AI}}=\frac{176}{176+43}=\frac{176}{219}=80.4\%$$

$$\text{NPV}_{\text{AI}}=\frac{969}{969+14}=\frac{969}{983}=98.6\%$$

and

$$\text{Accuracy}_{\text{AI}}=\frac{176+969}{1{,}202}=\frac{1{,}145}{1{,}202}=95.3\%$$

For radiologists:

$$\text{Sensitivity}_{\text{Rad}}=\frac{171}{171+19}=\frac{171}{190}=90.0\%$$

$$\text{Specificity}_{\text{Rad}}=\frac{1{,}003}{1{,}003+9}=\frac{1{,}003}{1{,}012}=99.1\%$$

$$\text{PPV}_{\text{Rad}}=\frac{171}{171+9}=\frac{171}{180}=95.0\%$$

$$\text{NPV}_{\text{Rad}}=\frac{1{,}003}{1{,}003+19}=\frac{1{,}003}{1{,}022}=98.1\%$$

and

$$\text{Accuracy}_{\text{Rad}}=\frac{171+1{,}003}{1{,}202}=\frac{1{,}174}{1{,}202}=97.7\%$$

These independently reconstructed conventional performance metrics reproduced the values reported by Ben Cheikh *et al.*, providing an internal consistency check for the reconstructed marginal and paired matrices.

The final values carried forward into Table 3 were therefore PNA=1.58% (19/1,202), NNA=3.99% (48/1,202), and NENA=-2.41% (-29/1,202). The corresponding reconstructed conventional performance metrics were 92.6% sensitivity, 95.8% specificity, 80.4% PPV, 98.6% NPV, and 95.3% accuracy for AI, and 90.0% sensitivity, 99.1% specificity, 95.0% PPV, 98.1% NPV, and 97.7% accuracy for the radiologists. These are the values reported in Table 3 of the present study.

### APPENDIX A3

#### Reconstruction of data from McKinney *et al*.

*McKinney et al.: UK and US screening mammography cohorts*

McKinney *et al*. provided paired confusion matrices comparing the AI system with human clinical decisions in Supplementary Table 1 of the original publication. Separate matrices were reported for reference-standard-positive and reference-standard-negative examinations for three comparisons: the UK first reader at 39 months; the UK consensus decision at 39 months; and the US first reader at 27 months. These published matrices provided sufficient same-case information to reconstruct the numbers of reference-standard-positive and reference-standard-negative examinations, disease prevalence, PNA, NNA, and NENA. They also allowed reconstruction of separate reference-standard confusion matrices for AI and for the corresponding first-reader or consensus decision, from which conventional diagnostic performance metrics were calculated.

For each comparison, the numbers of reference-standard-positive and reference-standard-negative examinations were first obtained by summing the four cells of the corresponding matrices in Supplementary Table 1. The total number of examinations and disease prevalence were then calculated. Within reference-standard-positive examinations, an AI TP/reader FN combination represented PNA, whereas an AI FN/reader TP combination represented NNA. Within reference-standard-negative examinations, an AI TN/reader FP combination represented PNA, whereas an AI FP/reader TN combination represented NNA. The paired matrices were subsequently collapsed according to AI and reader/consensus classifications to reconstruct conventional confusion matrices against the reference standard. The sensitivity, specificity, PPV, NPV, and accuracy were then calculated separately for AI and the corresponding human comparator.

**UK cohort: AI versus first reader**

*Reconstruction of cohort size and disease prevalence*

For the UK first-reader comparison, Supplementary Table 1 reported the following paired matrix among reference-standard-positive examinations:

| | First reader TP | First reader FN | Total |
|---|---|---|---|
| AI TP | 220 | 45 | 265 |
| AI FN | 34 | 108 | 142 |
| Total | 254 | 153 | 407 |

The number of reference-standard-positive examinations was therefore:

$$220+45+34+108=407$$

Among reference-standard-negative examinations, the paired matrix was:

| | First reader TN | First reader FP | Total |
|---|---|---|---|
| AI TN | 22,165 | 1,439 | 23,604 |
| AI FP | 1,175 | 336 | 1,511 |
| Total | 23,340 | 1,775 | 25,115 |

The number of reference-standard-negative examinations was therefore:

$$22{,}165+1{,}439+1{,}175+336=25{,}115$$

The total number of examinations was consequently:

$$Q=407+25{,}115=25{,}522$$

Disease prevalence was:

$$\text{Prevalence}=\frac{407}{25{,}522}=1.59\%$$

*Calculation of PNA, NNA, and NENA*

Among reference-standard-positive examinations, 45 examinations were AI TP/first-reader FN and therefore represented PNA, whereas 34 examinations were AI FN/first-reader TP and represented NNA. Among reference-standard-negative examinations, 1,439 examinations were AI TN/first-reader FP and represented PNA, whereas 1,175 examinations were AI FP/first-reader TN and represented NNA. The total number of PNA events was therefore:

$$P=45+1{,}439=1{,}484$$

and:

$$PNA=\frac{1{,}484}{25{,}522}=5.81\%$$

The total number of NNA events was:

$$N=34+1{,}175=1{,}209$$

and:

$$NNA=\frac{1{,}209}{25{,}522}=4.74\%$$

NENA was therefore:

$$NENA=\frac{P-N}{Q}$$
$$=\frac{1{,}484-1{,}209}{25{,}522}$$
$$=\frac{275}{25{,}522}=+1.08\%$$

Thus, in the UK first-reader comparison, AI was correct while the first reader was incorrect on 1,484 examinations, whereas the first reader was correct while AI was incorrect on 1,209 examinations, resulting in a net directional difference of 275 examinations in favor of AI.

*Reconstruction of conventional confusion matrices*

The paired matrices were collapsed according to AI classification to reconstruct the following AI-*versus*-reference-standard matrix:

| | Reference standard positive | Reference standard negative | Total |
|---|---|---|---|
| AI positive | 265 (TP) | 1,511 (FP) | 1,776 |
| AI negative | 142 (FN) | 23,604 (TN) | 23,746 |
| Total | 407 | 25,115 | 25,522 |

The same published matrices were collapsed according to the first-reader classification to reconstruct:

| | Reference standard positive | Reference standard negative | Total |
|---|---|---|---|
| First reader positive | 254 (TP) | 1,775 (FP) | 2,029 |
| First reader negative | 153 (FN) | 23,340 (TN) | 23,493 |
| Total | 407 | 25,115 | 25,522 |

For AI:

$$\text{Sensitivity}_{AI}=\frac{265}{407}=65.1\%$$

$$\text{Specificity}_{AI}=\frac{23{,}604}{25{,}115}=94.0\%$$

$$\text{PPV}_{AI}=\frac{265}{265+1{,}511}=14.9\%$$

$$\text{NPV}_{AI}=\frac{23{,}604}{23{,}604+142}=99.4\%$$

and

$$\text{Accuracy}_{\text{AI}}=\frac{265+23,604}{25,522}=93.5\%$$

For the first reader:

$$\text{Sensitivity}_{\text{Reader}}=\frac{254}{407}=62.4\%$$

$$\text{Specificity}_{\text{Reader}}=\frac{23,340}{25,115}=92.9\%$$

$$\text{PPV}_{\text{Reader}}=\frac{254}{254+1,775}=12.5\%$$

$$\text{NPV}_{\text{Reader}}=\frac{23,340}{23,340+153}=99.4\%$$

and

$$\text{Accuracy}_{\text{Reader}}=\frac{254+23,340}{25,522}=92.5\%$$

**UK cohort: AI *versus* consensus decision**

*Reconstruction of cohort size and disease prevalence*

For the UK consensus comparison, the paired matrix among reference-standard-positive examinations was:

| | Consensus TP | Consensus FN | Total |
|---|---|---|---|
| AI TP | 248 | 40 | 288 |
| AI FN | 31 | 95 | 126 |
| Total | 279 | 135 | 414 |

The number of reference-standard-positive examinations was therefore:

$$248+40+31+95=414$$

Among reference-standard-negative examinations, the paired matrix was:

| | Consensus TN | Consensus FP | Total |
|---|---|---|---|
| AI TN | 22,683 | 713 | 23,396 |
| AI FP | 1,803 | 243 | 2,046 |
| Total | 24,486 | 956 | 25,442 |

The number of reference-standard-negative examinations was therefore:

$$22{,}683+713+1{,}803+243=25{,}442$$

The total cohort size was:

$$Q=414+25{,}442=25{,}856$$

Disease prevalence was:

$$\text{Prevalence}=\frac{414}{25{,}856}=1.60\%$$

*Calculation of PNA, NNA, and NENA*

Among reference-standard-positive examinations, 40 AI TP/consensus FN examinations represented PNA, whereas 31 AI FN/consensus TP examinations represented NNA. Among reference-standard-negative examinations, 713 AI TN/consensus FP examinations represented PNA, whereas 1,803 AI FP/consensus TN examinations represented NNA. The total number of PNA events was therefore:

$$P=40+713=753$$

giving:

$$\text{PNA}=\frac{753}{25{,}856}=2.91\%$$

The total number of NNA events was:

$$N=31+1{,}803=1{,}834$$

giving:

$$\text{NNA}=\frac{1{,}834}{25{,}856}=7.09\%$$

NENA was therefore:

$$\mathrm{NENA}=\frac{753-1{,}834}{25{,}856}$$

$$=\frac{-1{,}081}{25{,}856}=-4.18\%$$

Thus, AI was correct while the consensus decision was incorrect on 753 examinations, whereas the consensus decision was correct while AI was incorrect on 1,834 examinations, resulting in a net directional difference of 1,081 examinations in favor of the consensus decision.

*Reconstruction of conventional confusion matrices*

Collapsing the paired matrices according to AI classification yielded:

| | Reference standard positive | Reference standard negative | Total |
|---|---|---|---|
| AI positive | 288 (TP) | 2,046 (FP) | 2,334 |
| AI negative | 126 (FN) | 23,396 (TN) | 23,522 |
| Total | 414 | 25,442 | 25,856 |

Collapsing the same matrices according to the consensus decision yielded:

| | Reference standard positive | Reference standard negative | Total |
|---|---|---|---|
| Consensus positive | 279 (TP) | 956 (FP) | 1,235 |
| Consensus negative | 135 (FN) | 24,486 (TN) | 24,621 |
| Total | 414 | 25,442 | 25,856 |

For AI:

$$\mathrm{Sensitivity}_{\mathrm{AI}}=\frac{288}{414}=69.6\%$$

$$\mathrm{Specificity}_{\mathrm{AI}}=\frac{23{,}396}{25{,}442}=92.0\%$$

$$\mathrm{PPV}_{\mathrm{AI}}=\frac{288}{288+2{,}046}=12.3\%$$

$$\mathrm{NPV}_{\mathrm{AI}}=\frac{23{,}396}{23{,}396+126}=99.5\%$$

and

$$\mathrm{Accuracy}_{\mathrm{AI}}=\frac{288+23{,}396}{25{,}856}=91.6\%$$

For the consensus decision:

$$\mathrm{Sensitivity}_{\mathrm{Consensus}}=\frac{279}{414}=67.4\%$$

$$\mathrm{Specificity}_{\mathrm{Consensus}}=\frac{24{,}486}{25{,}442}=96.2\%$$

$$\mathrm{PPV}_{\mathrm{Consensus}}=\frac{279}{279+956}=22.6\%$$

$$\mathrm{NPV}_{\mathrm{Consensus}}=\frac{24{,}486}{24{,}486+135}=99.5\%$$

and

$$\mathrm{Accuracy}_{\mathrm{Consensus}}=\frac{279+24{,}486}{25{,}856}=95.8\%$$

**US cohort: AI *versus* first reader**

*Reconstruction of cohort size and disease prevalence*

For the US first-reader comparison, the paired matrix among reference-standard-positive examinations was:

| | First reader TP | First reader FN | Total |
|---|---|---|---|
| AI TP | 197 | 121 | 318 |
| AI FN | 69 | 166 | 235 |
| Total | 266 | 287 | 553 |

The number of reference-standard-positive examinations was therefore:

$$197+121+69+166=553$$

Among reference-standard-negative examinations, the paired matrix was:

| | First reader TN | First reader FP | Total |
|---|---|---|---|
| AI TN | 1,282 | 482 | 1,764 |
| AI FP | 240 | 181 | 421 |
| Total | 1,522 | 663 | 2,185 |

The number of reference-standard-negative examinations was therefore:

$$1,282+482+240+181=2,185$$

The total cohort size was:

$$Q=553+2,185=2,738$$

Disease prevalence was:

$$\text{Prevalence}=\frac{553}{2,738}=20.20\%$$

*Calculation of PNA, NNA, and NENA*

Among reference-standard-positive examinations, 121 AI TP/first-reader FN examinations represented PNA, whereas 69 AI FN/first-reader TP examinations represented NNA. Among reference-standard-negative examinations, 482 AI TN/first-reader FP examinations represented PNA, whereas 240 AI FP/first-reader TN examinations represented NNA. The total number of PNA events was:

$$P=121+482=603$$

giving:

$$\text{PNA}=\frac{603}{2,738}=22.02\%$$

The total number of NNA events was:

$$N=69+240=309$$

giving:

$$\text{NNA}=\frac{309}{2,738}=11.29\%$$

NENA was therefore:

$$\text{NENA}=\frac{603\text{-}309}{2,738}$$

$$=\frac{294}{2,738}=+10.74\%$$

Thus, AI was correct while the first reader was incorrect on 603 examinations, whereas the first reader was correct while AI was incorrect on 309 examinations, resulting in a net directional difference of 294 examinations in favor of AI.

*Reconstruction of conventional confusion matrices*

Collapsing the paired matrices according to AI classification yielded:

| | Reference standard positive | Reference standard negative | Total |
|---|---|---|---|
| AI positive | 318 (TP) | 421 (FP) | 739 |
| AI negative | 235 (FN) | 1,764 (TN) | 1,999 |
| Total | 553 | 2,185 | 2,738 |

Collapsing the same matrices according to the first-reader classification yielded:

| | Reference standard positive | Reference standard negative | Total |
|---|---|---|---|
| First reader positive | 266 (TP) | 663 (FP) | 929 |
| First reader negative | 287 (FN) | 1,522 (TN) | 1,809 |
| Total | 553 | 2,185 | 2,738 |

For AI:

$$\text{Sensitivity}_{\text{AI}}=\frac{318}{553}=57.5\%$$

$$\text{Specificity}_{\text{AI}}=\frac{1{,}764}{2{,}185}=80.7\%$$

$$\text{PPV}_{\text{AI}}=\frac{318}{318+421}=43.0\%$$

$$\text{NPV}_{\text{AI}}=\frac{1{,}764}{1{,}764+235}=88.2\%$$

and

$$\text{Accuracy}_{\text{AI}}=\frac{318+1{,}764}{2{,}738}=76.0\%$$

For the first reader:

$$\text{Sensitivity}_{\text{Reader}}=\frac{266}{553}=48.1\%$$

$$\text{Specificity}_{\text{Reader}}=\frac{1{,}522}{2{,}185}=69.7\%$$

$$\text{PPV}_{\text{Reader}}=\frac{266}{266+663}=28.6\%$$

$$\text{NPV}_{\text{Reader}}=\frac{1{,}522}{1{,}522+287}=84.1\%$$

and

$$\text{Accuracy}_{\text{Reader}}=\frac{266+1{,}522}{2{,}738}=65.3\%$$

**Summary of reconstruction**

McKinney *et al.*'s Supplementary Table 1 provided all information required to reconstruct the cohort denominators, disease prevalence, directional non-agreement metrics, and conventional diagnostic performance metrics for all three comparisons. The reconstructed prevalence and directional metrics were:

| Comparison | Prevalence (%) | PNA (%) | NNA (%) | NENA (%) |
|---|---|---|---|---|
| UK: AI *vs.* first reader | 1.59 (407/25,522) | 5.81 (1,484/25,522) | 4.74 (1,209/25,522) | +1.08 (+275/25,522) |
| UK: AI *vs.* consensus | 1.60 (414/25,856) | 2.91 (753/25,856) | 7.09 (1,834/25,856) | -4.18 (-1,081/25,856) |
| US: AI *vs.* first reader | 20.20 (553/2,738) | 22.02 (603/2,738) | 11.29 (309/2,738) | +10.74 (+294/2,738) |

The conventional sensitivity, specificity, PPV, NPV, and accuracy values for AI and the corresponding first-reader or consensus decision were independently calculated from the reconstructed reference-standard confusion matrices. This provided an internal consistency check of the extraction and reconstruction procedure. The reconstructed PNA, NNA, NENA, and conventional performance metrics were subsequently used in Table 3 of the present study.

**APPENDIX A4**

**Reconstruction of data from Goh *et al*.**

*Goh et al.: mammography interpreted by radiologists of varying experience*

Goh *et al*. included 500 women, of whom 250 had malignant findings and 250 had nonmalignant findings, corresponding to a patient-level disease prevalence of:

$$\frac{250}{500}=50.0\%$$

The study included 17 radiologists stratified by experience: four consultants, four senior residents, and nine junior residents. Each radiologist interpreted all 500 examinations. Accordingly, analyses of AI-radiologist non-agreement were reconstructed at the paired reading level rather than at the patient level.

This yielded:

$$4\times500=2{,}000$$

paired readings for consultants,

$$4\times500=2{,}000$$

paired readings for senior residents, and

$$9\times500=4{,}500$$

paired readings for junior residents.

Because the study population contained 250 malignant and 250 nonmalignant examinations, these denominators comprised 1,000 reference-standard-positive and 1,000 reference-standard-negative readings for consultants and senior residents, and

2,250 reference-standard-positive and 2,250 reference-standard-negative readings for junior residents.

Goh *et al*. reported the AI-alone sensitivity and specificity in Figure 1 and provided, in Table 3, exact counts for four directions of AI-radiologist discordances: AI correct/radiologist false negative, AI correct/radiologist false positive, radiologist correct/AI false negative, and radiologist correct/AI false positive. These data were sufficient to reconstruct the paired AI-radiologist matrices and the corresponding radiologist marginal confusion matrices. The reconstruction used the exact AI counts implied by the 250 malignant and 250 nonmalignant examinations together with the exact discordance counts reported in Table 3.

*AI-alone reference-standard confusion matrix*

The AI system was evaluated on the same 250 malignant and 250 nonmalignant examinations. The reported AI sensitivity was 79.6%. With 250 reference-standard-positive examinations, this corresponds uniquely to:

$$TP_{AI}=0.796\times250=199$$

and:

$$FN_{AI}=250-199=51$$

The reported AI specificity was 90.8%. With 250 reference-standard-negative examinations, this corresponds uniquely to:

$$TN_{AI}=0.908\times250=227$$

and:

$$FP_{AI}=250-227=23$$

The resulting patient-level AI confusion matrix was therefore:

| | Reference standard positive | Reference standard negative | Total |
|---|---|---|---|
| AI positive | 199 (TP) | 23 (FP) | 222 |
| AI negative | 51 (FN) | 227 (TN) | 278 |
| Total | 250 | 250 | 500 |

The corresponding conventional performance metrics were:

$$Sensitivity_{AI}=\frac{199}{250}=79.6\%$$

$$Specificity_{AI}=\frac{227}{250}=90.8\%$$

$$PPV_{AI}=\frac{199}{199+23}=\frac{199}{222}=89.6\%$$

$$NPV_{AI}=\frac{227}{227+51}=\frac{227}{278}=81.7\%$$

and:

$$Accuracy_{AI}=\frac{199+227}{500}=\frac{426}{500}=85.2\%$$

Because every radiologist interpreted the same 500 examinations, these AI counts were replicated according to the number of radiologists in each experience group when constructing the paired reading-level matrices.

**Consultants**

*Reconstruction of reading-level denominators*

Four consultants each interpreted 500 examinations, yielding:

$$4\times500=2{,}000$$

paired AI-consultant readings.

Of these:

$$4 \times 250 = 1{,}000$$

were reference-standard positive and 1,000 were reference-standard negative.

Replicating the AI-alone confusion matrix across four consultants yielded:

$$TP_{AI} = 199 \times 4 = 796$$
$$FN_{AI} = 51 \times 4 = 204$$
$$TN_{AI} = 227 \times 4 = 908$$

and:

$$FP_{AI} = 23 \times 4 = 92$$

*Reconstruction among reference-standard-positive readings*

Table 3 reported 157 readings in which AI was correct and the consultant was false negative, and 66 readings in which the consultant was correct and AI was false negative. These corresponded to:

- AI TP / consultant FN = 157
- AI FN / consultant TP = 66

Because AI had 796 TP readings:

$$\text{AI TP/consultant TP} = 796 - 157 = 639$$

Because AI had 204 FN readings:

$$\text{AI FN/consultant FN} = 204 - 66 = 138$$

The paired matrix among the 1,000 reference-standard-positive readings was therefore:

| | Consultant TP | Consultant FN | Total |
|---|---|---|---|
| AI TP | 639 | 157 | 796 |
| AI FN | 66 | 138 | 204 |
| Total | 705 | 295 | 1,000 |

The reconstructed consultant sensitivity was:

$$\text{Sensitivity}_{\text{consultant}}=\frac{705}{1,000}=70.5\%$$

*Reconstruction among reference-standard-negative readings*

Table 3 reported 17 readings in which AI was correct and the consultant was false positive, and 79 readings in which the consultant was correct and AI was false positive. These corresponded to:

- AI TN / consultant FP = 17
- AI FP / consultant TN = 79

The remaining concordant cells were therefore:

$$\text{AI TN/consultant TN}=908-17=891$$

and:

$$\text{AI FP/consultant FP}=92-79=13$$

The paired matrix among the 1,000 reference-standard-negative readings was:

| | Consultant TN | Consultant FP | Total |
|---|---|---|---|
| AI TN | 891 | 17 | 908 |
| AI FP | 79 | 13 | 92 |
| Total | 970 | 30 | 1,000 |

The reconstructed consultant specificity was:

$$\text{Specificity}_{\text{consultant}}=\frac{970}{1,000}=97.0\%$$

*Calculation of PNA, NNA, and NENA*

PNA comprised readings in which AI was correct and the consultant was incorrect:

$$P_{\text{consultant}}=157+17=174$$

and:

$$PNA_{consultant}=\frac{174}{2,000}=8.70\%$$

NNA comprised readings in which the consultant was correct and AI was incorrect:

$$N_{consultant}=66+79=145$$

and:

$$NNA_{consultant}=\frac{145}{2,000}=7.25\%.$$

NENA was:

$$NENA_{consultant}=PNA-NNA$$
$$=\frac{174-145}{2,000}$$
$$=\frac{29}{2,000}=+1.45\%$$

*Reconstruction of conventional consultant performance*

The reconstructed consultant-*versus*-reference-standard matrix was:

| | Reference standard positive | Reference standard negative | Total |
|---|---|---|---|
| Consultant positive | 705 (TP) | 30 (FP) | 735 |
| Consultant negative | 295 (FN) | 970 (TN) | 1,265 |
| Total | 1,000 | 1,000 | 2,000 |

Thus:

$$Sensitivity=\frac{705}{1,000}=70.5\%$$

$$Specificity=\frac{970}{1,000}=97.0\%$$

$$PPV=\frac{705}{735}=95.9\%$$

$$NPV=\frac{970}{1,265}=76.7\%$$

and:

$$\text{Accuracy}=\frac{705+970}{2{,}000}=\frac{1{,}675}{2{,}000}=83.75\%$$

**Senior residents**

*Reconstruction of reading-level denominators*

Four senior residents each interpreted 500 examinations, yielding:

$$4\times500=2{,}000$$

paired readings, comprising 1,000 reference-standard-positive and 1,000 reference-standard-negative readings.

The replicated AI totals were again:

796 TP, 204 FN, 908 TN, 92 FP

*Reconstruction among reference-standard-positive readings*

Table 3 reported 215 readings in which AI was correct and the senior resident was false negative, and 60 readings in which the senior resident was correct and AI was false negative.

Thus:

$$\text{AI TP/senior TP}=796-215=581$$

and:

$$\text{AI FN/senior FN}=204-60=144$$

The paired matrix was:

| | Senior resident TP | Senior resident FN | Total |
|---|---|---|---|
| AI TP | 581 | 215 | 796 |
| AI FN | 60 | 144 | 204 |
| Total | 641 | 359 | 1,000 |

The reconstructed senior-resident sensitivity was:

$$\text{Sensitivity}_{\text{senior}}=\frac{641}{1,000}=64.1\%$$

*Reconstruction among reference-standard-negative readings*

Table 3 reported 23 readings in which AI was correct and the senior resident was false positive, and 82 readings in which the senior resident was correct and AI was false positive. The concordant cells were therefore:

$$\text{AI TN/senior TN}=908-23=885$$

and:

$$\text{AI FP/senior FP}=92-82=10$$

The paired matrix was:

| | Senior resident TN | Senior resident FP | Total |
|---|---|---|---|
| AI TN | 885 | 23 | 908 |
| AI FP | 82 | 10 | 92 |
| Total | 967 | 33 | 1,000 |

The reconstructed senior-resident specificity was:

$$\text{Specificity}_{\text{senior}}=\frac{967}{1,000}=96.7\%$$

*Calculation of PNA, NNA, and NENA*

PNA was:

$$P_{\text{senior}}=215+23=238$$

$$\text{PNA}_{\text{senior}}=\frac{238}{2,000}=11.90\%$$

NNA was:

$$N_{senior}=60+82=142$$

$$NNA_{senior}=\frac{142}{2,000}=7.10\%$$

NENA was:

$$NENA_{senior}=\frac{238-142}{2,000}$$

$$=\frac{96}{2,000}=+4.80\%$$

*Reconstruction of conventional senior-resident performance*

The reconstructed senior-resident-*versus*-reference-standard matrix was:

| | Reference standard positive | Reference standard negative | Total |
|---|---|---|---|
| Senior resident positive | 641 (TP) | 33 (FP) | 674 |
| Senior resident negative | 359 (FN) | 967 (TN) | 1,326 |
| Total | 1,000 | 1,000 | 2,000 |

Thus:

$$Sensitivity=\frac{641}{1,000}=64.1\%$$

$$Specificity=\frac{967}{1,000}=96.7\%$$

$$PPV=\frac{641}{674}=95.1\%$$

$$NPV=\frac{967}{1,326}=72.9\%$$

and:

$$Accuracy=\frac{641+967}{2,000}=\frac{1,608}{2,000}=80.4\%.$$

**Junior residents**

*Reconstruction of reading-level denominators*

Nine junior residents each interpreted 500 examinations, yielding:

$$9 \times 500 = 4{,}500$$

paired readings.

These comprised:

$$9 \times 250 = 2{,}250$$

reference-standard-positive readings and 2,250 reference-standard-negative readings.

Replicating the exact patient-level AI counts across nine junior residents yielded:

$$TP_{AI} = 199 \times 9 = 1{,}791$$
$$FN_{AI} = 51 \times 9 = 459$$
$$TN_{AI} = 227 \times 9 = 2{,}043$$

and:

$$FP_{AI} = 23 \times 9 = 207$$

*Reconstruction among reference-standard-positive readings*

Table 3 reported 502 readings in which AI was correct and the junior resident was false negative, and 97 readings in which the junior resident was correct and AI was false negative.

Thus:

$$\text{AI TP/junior TP} = 1{,}791 - 502 = 1{,}289$$

and:

$$\text{AI FN/junior FN} = 459 - 97 = 362$$

The paired matrix was therefore:

| | Junior resident TP | Junior resident FN | Total |
|---|---|---|---|
| AI TP | 1,289 | 502 | 1,791 |
| AI FN | 97 | 362 | 459 |
| Total | 1,386 | 864 | 2,250 |

The reconstructed junior-resident sensitivity was:

$$\text{Sensitivity}_{\text{junior}} = \frac{1{,}386}{2{,}250} = 61.6\%$$

*Reconstruction among reference-standard-negative readings*

Table 3 reported 58 readings in which AI was correct and the junior resident was false positive, and 181 readings in which the junior resident was correct and AI was false positive. These corresponded to:

- AI TN / junior resident FP = 58
- AI FP / junior resident TN = 181

Because the replicated AI marginal counts contained exactly 2,043 TN readings:

$$\text{AI TN/junior TN} = 2{,}043 - 58 = 1{,}985$$

Because the replicated AI marginal counts contained exactly 207 FP readings:

$$\text{AI FP/junior FP} = 207 - 181 = 26$$

The paired matrix was therefore uniquely reconstructed as:

| | Junior resident TN | Junior resident FP | Total |
|---|---|---|---|
| AI TN | 1,985 | 58 | 2,043 |
| AI FP | 181 | 26 | 207 |
| Total | 2,166 | 84 | 2,250 |

The reconstructed junior-resident specificity was:

$$\text{Specificity}_{\text{junior}} = \frac{2{,}166}{2{,}250} = 96.27\%$$

Thus, when the exact AI marginal counts were combined with the exact Table 3 discordance counts, the junior-resident negative-case reconstruction was uniquely determined as 2,166 TN and 84 FP readings. This exact reconstruction yielded a junior-resident accuracy of 78.9%, whereas Figure 1 of the original publication displays an accuracy of 79.0%. A reconstruction of 2,167 TN and 83 FP would have reproduced the displayed 79.0% accuracy but would have been incompatible by one reading with the exact replicated AI marginal counts and the exact discordance counts reported in Table 3. The discrepancy therefore represented a minor internal reporting inconsistency in the original publication rather than ambiguity in the reconstruction used in the present analysis. Importantly, this one-reading discrepancy did not affect any of the directional discordance counts reported in Table 3 and therefore did not affect PNA, NNA, or NENA.

*Calculation of PNA, NNA, and NENA*

PNA comprised the two Table 3 categories in which AI was correct and the junior resident was incorrect:

$$P_{junior}=502+58=560$$

and:

$$PNA_{junior}=\frac{560}{4,500}=12.44\%$$

NNA comprised the two categories in which the junior resident was correct and AI was incorrect:

$$N_{junior}=97+181=278$$

and:

$$NNA_{junior}=\frac{278}{4,500}=6.18\%$$

NENA was therefore:

$$NENA_{junior}=\frac{560-278}{4,500}$$

$$=\frac{282}{4,500}=+6.27\%$$

*Reconstruction of conventional junior-resident performance*

The reconstructed junior-resident-*versus*-reference-standard matrix was:

| | Reference standard positive | Reference standard negative | Total |
|---|---|---|---|
| Junior resident positive | 1,386 (TP) | 84 (FP) | 1,470 |
| Junior resident negative | 864 (FN) | 2,166 (TN) | 3,030 |
| Total | 2,250 | 2,250 | 4,500 |

Thus:

$$Sensitivity=\frac{1,386}{2,250}=61.6\%$$

$$Specificity=\frac{2,166}{2,250}=96.3\%$$

$$PPV=\frac{1,386}{1,386+84}=\frac{1,386}{1,470}=94.3\%$$

$$NPV=\frac{2,166}{2,166+864}=\frac{2,166}{3,030}=71.5\%$$

and:

$$Accuracy=\frac{1,386+2,166}{4,500}=\frac{3,552}{4,500}=78.93\%$$

**Summary of reconstruction**

Goh *et al*. included 500 women, with a patient-level disease prevalence of 50.0% (250/500). Because each examination was interpreted by multiple radiologists, the directional AI-radiologist comparisons were reconstructed at the paired reading level, yielding denominators of 2,000 readings for consultants, 2,000 for senior residents, and 4,500 for junior residents. The four exact discordance categories reported in Table 3 directly determined the PNA and NNA counts. NENA was calculated as PNA minus NNA. The reconstructed directional metrics were:

| Comparison | PNA (%) | NNA (%) | NENA (%) |
|---|---|---|---|
| AI *vs.* 4 consultants | 8.70 (174/2,000) | 7.25 (145/2,000) | +1.45 (+29/2,000) |
| AI *vs*. 4 senior residents | 11.90 (238/2,000) | 7.10 (142/2,000) | +4.80 (+96/2,000) |
| AI *vs*. 9 junior residents | 12.44 (560/4,500) | 6.18 (278/4,500) | +6.27 (+282/4,500) |

The reconstructed conventional performance metrics were:

| Reader group | Sensitivity (%) | Specificity (%) | PPV (%) | NPV (%) | Accuracy (%) |
|---|---|---|---|---|---|
| AI | 79.6 | 90.8 | 89.6 | 81.7 | 85.2 |
| Consultants | 70.5 | 97.0 | 95.9 | 76.7 | 83.8 |
| Senior residents | 64.1 | 96.7 | 95.1 | 72.9 | 80.4 |
| Junior residents | 61.6 | 96.3 | 94.3 | 71.5 | 78.9 |

The AI-alone confusion matrix was determined by the reported sensitivity and specificity because these percentages corresponded exactly to integer counts among 250 malignant and 250 nonmalignant examinations. Replication of these AI counts across the radiologists in each experience group, together with the exact Table 3 discordance counts, determined the paired AI-radiologist matrices and radiologist marginal counts.

For junior residents, this reconstruction yielded 1,386 TP, 864 FN, 2,166 TN, and 84 FP readings. These counts reproduce the reported sensitivity of 61.6% and specificity of

96.3% and yield an accuracy of 78.9%. The 79.0% accuracy displayed in Figure 1 would require one additional TN reading and one fewer FP reading, which is incompatible with the exact AI marginal and Table 3 discordance counts. This one-reading discrepancy in the original publication does not affect the reported discordance counts or the resulting PNA, NNA, and NENA values. Accordingly, the values carried forward into the applicability analysis were PNA=8.70%, NNA=7.25%, and NENA=+1.45% for the consultants; PNA=11.90%, NNA=7.10%, and NENA=+4.80% for the senior residents; and PNA=12.44%, NNA=6.18%, and NENA=+6.27% for the junior residents.

**APPENDIX A5**

**Illustrative scenarios: case-level reconstruction, confidence intervals, and effect of cohort size**

The three illustrative scenarios were specified as cohorts of 1,000 examinations, of which 400 were reference-standard positive and 600 reference-standard negative. Table A5.1 gives the paired AI-radiologist classification of every examination, from which all values in Table 2 and Figure 1 derive. Among reference-standard-positive examinations, AI true positive/radiologist false negative represents PNA and AI false negative/radiologist true positive represents NNA; among reference-standard-negative examinations, AI true negative/radiologist false positive represents PNA and AI false positive/radiologist true negative represents NNA.

**Table A5.1. Paired AI-radiologist classifications in illustrative scenarios (Q=1,000).**

| Scenario | Reference standard | AI and radiologist correct | AI correct, radiologist incorrect (PNA) | Radiologist correct, AI incorrect (NNA) | AI and radiologist incorrect | Total |
|---|---|---|---|---|---|---|
| 1 | Positive | 290 | 30 | 0 | 80 | 400 |
| 1 | Negative | 490 | 30 | 10 | 70 | 600 |
| 2 | Positive | 190 | 130 | 70 | 10 | 400 |
| 2 | Negative | 310 | 70 | 80 | 140 | 600 |
| 3 | Positive | 230 | 90 | 30 | 50 | 400 |
| 3 | Negative | 460 | 0 | 90 | 50 | 600 |

Confidence intervals and the reclassification FI were obtained as described in Appendix A1, using 10,000 multinomial bootstrap resamples (random seed 2026). Results are given in Table A5.2. The percentile limits obtained by exact enumeration of the multinomial bootstrap distribution (the limit of an infinite number of resamples) differed by at most 0.1 percentage point, and the reclassification FI was identical.

**Table A5.2. Directional metrics with 95% bootstrap confidence intervals and reclassification fragility index (Q=1,000).**

| Scenario | PNA (%)<br>(95% CI) | NNA (%)<br>(95% CI) | NENA (%)<br>(95% CI) | FI (%) |
|---|---|---|---|---|
| 1 | 6.0<br>(4.6 to 7.5) | 1.0<br>(0.4 to 1.7) | +5.0<br>(+3.4 to +6.6) | 1.70<br>(17/1,000) |
| 2 | 20.0<br>(17.5 to 22.5) | 15.0<br>(12.9 to 17.2) | +5.0<br>(+1.3 to +8.7) | 0.70<br>(7/1,000) |
| 3 | 9.0<br>(7.3 to 10.8) | 12.0<br>(10.0 to 14.0) | -3.0<br>(-5.9 to -0.2) | 0.10<br>(1/1,000) |

To show which values depend on cohort size, the three scenarios were recalculated with identical proportions for Q ranging from 100 to 10,000 (Table A5.3). PNA, NNA, NENA, sensitivity, specificity, PPV, NPV, and accuracy were identical at every cohort size; only the confidence intervals and the reclassification FI changed. At Q=100, the 95% confidence interval for NENA included 0 in all three scenarios and the reclassification FI was undefined. Among cohort sizes for which the scenario counts were integers, the confidence interval first excluded 0 at approximately Q=200, Q=560, and Q=1,000 for scenarios 1, 2, and 3, respectively. For a given NENA, the reclassification FI expressed as a percentage of Q increased with Q toward approximately half the absolute value of NENA, the value of the point-estimate-based count described in Appendix A1 (2.5% for scenarios 1 and 2, and 1.5% for scenario 3). The reclassification FI therefore reflects both the magnitude of NENA and the sample size on which it is estimated.

**Table A5.3. 95% bootstrap confidence interval for NENA and reclassification fragility index by cohort size, at constant proportions.**

| Q | Scenario 1 (NENA +5%): 95% CI (%) | Scenario 1: FI (%) | Scenario 2 (NENA +5%): 95% CI (%) | Scenario 2: FI (%) | Scenario 3 (NENA -3%): 95% CI (%) | Scenario 3: FI (%) |
|---|---|---|---|---|---|---|
| 100 | 0.00 to +11.00 | not applicable | -7.00 to +17.00 | not applicable | -12.00 to +6.00 | not applicable |
| 200 | +1.50 to +9.00 | 1.00 (2/200) | -3.00 to +13.00 | not applicable | -9.50 to +3.50 | not applicable |
| 500 | +2.80 to +7.40 | 1.40 (7/500) | -0.40 to +10.20 | not applicable | -7.00 to +1.00 | not applicable |
| 1,000 | +3.40 to +6.60 | 1.70 (17/1,000) | +1.30 to +8.70 | 0.70 (7/1,000) | -5.90 to -0.20 | 0.10 (1/1,000) |
| 2,000 | +3.85 to +6.15 | 1.90 (38/2,000) | +2.45 to +7.55 | 1.20 (24/2,000) | -5.05 to -1.00 | 0.50 (10/2,000) |
| 10,000 | +4.49 to +5.52 | 2.24 (224/10,000) | +3.83 to +6.14 | 1.92 (192/10,000) | -3.93 to -2.10 | 1.05 (105/10,000) |

**APPENDIX A6**

**Bootstrap confidence intervals and reclassification fragility index for the applicability studies**

For each of the seven applicability comparisons, the counts P, N, and Q reconstructed in Appendices A2 to A4 were entered into the procedure described in Appendix A1. Q assessments were resampled with replacement 10,000 times from three categories (PNA, NNA, and concordant) with probabilities P/Q, N/Q, and 1-(P+N)/Q; the 2.5th and 97.5th percentiles of the resampled PNA, NNA, and NENA formed the 95% confidence intervals. The random seed (2026) was reset before each interval so that every value is reproducible. When the 95% confidence interval for NENA excluded 0, assessments were reclassified from the majority to the minority discordant category, and the reclassification FI was the smallest number of reclassifications at which the recalculated interval included 0, located by binary search. The R implementation is given in Appendix A7.

**Table A6.1. Directional metrics with 95% bootstrap confidence intervals and reclassification fragility index for the applicability studies.**

| Comparison | Q | P | N | PNA (%) (95% CI) | NNA (%) (95% CI) | NENA (%) (95% CI) | FI, n (% of Q) |
|---|---|---|---|---|---|---|---|
| Ben Cheikh *et al.*: AI *vs*. radiologist | 1,202 | 19 | 48 | 1.58 (0.92 to 2.33) | 3.99 (2.91 to 5.16) | -2.41 (-3.74 to -1.08) | 7 (0.58) |
| McKinney *et al.* (UK): AI *vs*. first reader | 25,522 | 1,484 | 1,209 | 5.81 (5.52 to 6.10) | 4.74 (4.48 to 4.99) | +1.08 (+0.68 to +1.47) | 87 (0.34) |
| McKinney *et al.* (UK): AI *vs*. consensus decision | 25,856 | 753 | 1,834 | 2.91 (2.71 to 3.12) | 7.09 (6.78 to 7.41) | -4.18 (-4.57 to -3.80) | 491 (1.90) |
| McKinney *et al.* (US): AI *vs*. first reader | 2,738 | 603 | 309 | 22.02 (20.49 to 23.56) | 11.29 (10.12 to 12.49) | +10.74 (+8.62 to +12.82) | 118 (4.31) |
| Goh *et al*.: AI *vs*. 4 consultants | 2,000 | 174 | 145 | 8.70 (7.45 to 10.00) | 7.25 (6.15 to 8.40) | +1.45 (-0.35 to +3.20) | Not applicable |
| Goh *et al*.: AI *vs*. 4 senior residents | 2,000 | 238 | 142 | 11.90 (10.50 to 13.30) | 7.10 (6.00 to 8.25) | +4.80 (+2.90 to +6.70) | 29 (1.45) |
| Goh *et al*.: AI *vs*. 9 junior residents | 4,500 | 560 | 278 | 12.44 (11.49 to 13.42) | 6.18 (5.49 to 6.89) | +6.27 (+5.04 to +7.51) | 113 (2.51) |

The 95% confidence interval for NENA excluded 0 in 6 of the 7 comparisons, and the reclassification FI ranged from 0.34% (87/25,522 examinations; McKinney *et al.* (UK): AI *vs*. first reader) to 4.31% (118/2,738; McKinney *et al.* (US): AI *vs*. first reader). For the Goh *et al*. comparison against consultants, the interval included 0 and the reclassification FI was not applicable.

Because percentile limits were estimated from a finite number of resamples, they depended slightly on the random seed. Across 50 seeds, the confidence limits of NENA varied by at most 0.15 percentage point and the reclassification FI by at most three examinations (McKinney *et al*., UK first reader, 85 to 88), without changing whether any interval included 0. For the Goh *et al*. consultant comparison, the lower limit ranged from -0.35% to -0.25%. In Goh *et al*., every radiologist read all 500 examinations, so the reading-level denominators (2,000 and 4,500) contained repeated readings of the same examinations. The bootstrap treated each reading as independent (Appendix A1); for these three comparisons, the confidence intervals were therefore likely too narrow and the reclassification FI too large.

### APPENDIX A7

#### R code for the bootstrap confidence intervals and reclassification fragility index

The script below uses base R only and was run with R version 4.6.1. It reproduces every bootstrap confidence interval and the reclassification FI reported in the Results section, Table 2, and Appendices A1, A5, and A6. Each comparison requires only the counts P (AI-platform correct, radiologist incorrect), N (radiologist correct, AI-platform incorrect), and Q (total assessments). The function bootstrap_ci() returns percentile intervals for PNA, NNA, and NENA, and fragility_index() returns the reclassification FI as a number of examinations, or NA when the baseline interval for NENA already includes 0.

```
# Positive, negative and net non-agreement (PNA, NNA, NENA):
# multinomial bootstrap confidence intervals and reclassification fragility index (FI).
# Base R only. Inputs per comparison: P (AI right, radiologist wrong),
# N (radiologist right, AI wrong) and Q (total assessments).

B    <- 10000   # bootstrap resamples
SEED <- 2026    # reset before every interval, so results are reproducible
ALPHA <- 0.05

# Resample Q assessments from three categories (PNA, NNA, concordant) and
# return the percentile intervals of PNA, NNA and NENA.
bootstrap_ci <- function(P, N, Q, B_ = B, seed = SEED) {
  set.seed(seed)
  x <- rmultinom(B_, Q, c(P, N, Q - P - N) / Q)
  probs <- c(ALPHA / 2, 1 - ALPHA / 2)
  list(PNA  = quantile(x[1, ] / Q, probs, names = FALSE),
       NNA  = quantile(x[2, ] / Q, probs, names = FALSE),
       NENA = quantile((x[1, ] - x[2, ]) / Q, probs, names = FALSE))
}

includes_zero <- function(ci) ci[1] <= 0 && ci[2] >= 0

# Minimum number of assessments reclassified from the majority to the
# minority discordant category for the 95% CI of NENA to include zero.
# Returns NA when the baseline interval already includes zero (FI not applicable).
fragility_index <- function(P, N, Q) {
  if (includes_zero(bootstrap_ci(P, N, Q)$NENA)) return(NA_integer_)
  s  <- sign(P - N)
  lo <- 0             # interval excludes zero
  hi <- abs(P - N)    # P and N swapped past equality: interval includes zero
  # Binary search; inclusion of zero is monotone in the number reclassified.
  while (hi - lo > 1) {
    mid <- (lo + hi) %/% 2
    ci  <- bootstrap_ci(P - s * mid, N + s * mid, Q)$NENA
    if (includes_zero(ci)) hi <- mid else lo <- mid
  }
  hi
}

fmt_pct <- function(x, digits = 2, sign = FALSE)
  sprintf(paste0("%", if (sign) "+" else "", ".", digits, "f%%"), 100 * x)

fmt_ci <- function(est, ci, digits, sign = FALSE)
  sprintf("%s (%s to %s)", fmt_pct(est, digits, sign),
          fmt_pct(ci[1], digits, sign), fmt_pct(ci[2], digits, sign))

summarise_comparison <- function(label, P, N, Q, digits = 2) {
  ci <- bootstrap_ci(P, N, Q)
  fi <- fragility_index(P, N, Q)
  data.frame(
    comparison = label, Q = Q, P = P, N = N,
    PNA    = fmt_ci(P / Q, ci$PNA, digits),
    NNA    = fmt_ci(N / Q, ci$NNA, digits),
    NENA   = fmt_ci((P - N) / Q, ci$NENA, digits, sign = TRUE),
    FI_n   = fi,
    FI_pct = if (is.na(fi)) NA_character_ else fmt_pct(fi / Q, 2),
    stringsAsFactors = FALSE)
}

# ---- Applicability studies (Appendices A2-A4, Table 3) ----------------------
studies <- data.frame(
  comparison = c("Ben Cheikh et al.: AI vs. radiologist",
                 "McKinney et al., UK: AI vs. first reader",
                 "McKinney et al., UK: AI vs. consensus decision",
                 "McKinney et al., US: AI vs. first reader",
                 "Goh et al.: AI vs. 4 consultants",
                 "Goh et al.: AI vs. 4 senior residents",
                 "Goh et al.: AI vs. 9 junior residents"),
  P = c(19, 1484, 753, 603, 174, 238, 560),
  N = c(48, 1209, 1834, 309, 145, 142, 278),
  Q = c(1202, 25522, 25856, 2738, 2000, 2000, 4500))
```

```
summarise_all <- function(d, ...)
  do.call(rbind, Map(summarise_comparison, d$comparison, d$P, d$N, d$Q, ...))

applicability <- summarise_all(studies)

# ---- Illustrative scenarios, Q = 1,000 (Table 2, Appendix A5) ----------------
scenarios <- data.frame(comparison = paste("Scenario", 1:3),
                        P = c(60, 200, 90), N = c(10, 150, 120), Q = 1000)
illustrative <- summarise_all(scenarios, digits = 1)

# ---- Effect of cohort size at constant proportions (Appendix A5) ------------
by_q <- do.call(rbind, lapply(c(100, 200, 500, 1000, 2000, 10000), function(q)
  summarise_all(transform(scenarios, P = P * q / Q, N = N * q / Q, Q = q))))

out_dir <- "output"
dir.create(out_dir, showWarnings = FALSE)
save_csv <- function(x, name) write.csv(x, file.path(out_dir, name), row.names = FALSE)
save_csv(applicability, "applicability_bootstrap_fi.csv")
save_csv(illustrative,  "illustrative_q1000_bootstrap_fi.csv")
save_csv(by_q,          "illustrative_by_q_bootstrap_fi.csv")

options(width = 200)
print(applicability, row.names = FALSE)
print(illustrative, row.names = FALSE)
print(by_q[, c("comparison", "Q", "NENA", "FI_n", "FI_pct")], row.names = FALSE)
```
